# Gender differences in the distribution of children’s physical activity: evidence from nine countries

**DOI:** 10.1101/2023.04.17.23288558

**Authors:** Luke Kretschmer, Gul Deniz Salali, Lars Bo Andersen, Pedro C Hallal, Kate Northstone, Luís B. Sardinha, Mark Dyble, David Bann, International Children’s Accelerometry Database (ICAD) Collaborators

## Abstract

**Background:** Physical activity in childhood is thought to influences health and development. Previous studies have found that boys are typically more active than girls, yet the focus has largely been on differences in average levels or proportions above a threshold rather than the full distribution of activity across all intensities. We thus examined differences in the distribution of physical activity between girls and boys in a multi-national sample of children.

**Methods:** We used the harmonised International Children Accelerometery Database (ICAD), including waist-worn accelerometery data from 15,461 individuals (Boys: 48.3%) from 9 countries. Employing Generalised Additive Models of Location, Shape, and Scale (GAMLSS) we investigated gender differences in the distribution of individuals, including comparisons of variability (SD) and average physical activity levels (mean and median) and skewness. We conducted this analysis for each activity intensity (Sedentary, Light, and Moderate-to-Vigorous (MVPA)) and a summary measure (counts per minute (CPM)).

**Results:** Sizable gender differences in the distribution of activity were found for moderate to vigorous activity and counts per minute, with boys having higher average levels (38% higher mean volumes of MVPA, 20% higher CPM), yet substantially more between-person variability (30% higher standard deviation (SD) for MVPA, 17% higher SD for CPM); boys’ distributions were less positively skewed than girls. Conversely, there was little to no difference between girls and boys in the distribution of sedentary or light-intensity activity.

**Conclusions:** Inequality in activity between girls and boys was driven by MVPA. The higher mean volumes of MVPA in boys occurred alongside greater variability. This suggests a need to consider the underlying distribution of activity in future research; for example, interventions which target gender inequality in MVPA may inadvertently lead to increased inequality within girls.

## Introduction

Physical activity levels during childhood and adolescence have implications for health and development throughout the lifecourse.^1^ Low levels of activity in childhood have been linked to a series of unfavourable outcomes: higher incidence of infectious^2^ and chronic disease,^3–8^ poorer mental health outcomes,^3,9^ lower cognitive function and school performance,^9,10^ and delayed physical development.^3,7,8,10–15^ Gender is frequently observed to be a correlate of objectively measured physical activity in youth samples,^16,17^ with boys on average typically undertaking more activity than girls, with the effect size relatively stable across ages.^18–25^ Since childhood activity levels tend to track into later life,^26^ such differences may have lasting implications for gender disparities in subsequent health.^27^

Understanding the distribution of individuals across all active behaviours could help to better understand causes of gender differences in activity profiles However, such an approach is underutilised. ^28^ Research to date has largely focussed on comparing summary measures of physical activity (frequently average counts^29,30^ or MVPA^1,5,8,17,19,23^) with little research examining the distribution of activity across individuals. One identified paper investigated the Gini (an index of inequality for an outcome) of activity between countries, but did not examine gender.^28^ Analysing the full distribution of activity across all intensities, drivers of differences between girls and boys may be better understood, furthering an understanding of whether differences are due to a whole population shift, or owes to a subset skewing the sample.

To address this gap, the present research explores the full distribution of activity using Generalised Additive Models of Location Shape and Scale (GAMLSS) which allows for comparisons between medians, standard deviations and skews in addition to the mean.^31,32^ This analysis is repeated for the mean intensity of activity and each intensity threshold. Given the observed differences between girls and boys in volumes of MVPA, similar differences in the mean should be observed here. If this difference emerges due to volitional activity, such as sport or active play with a larger subset of one gender undertaking such activities,^33^ it may result in that gender having a wider distribution of activity and more skew. For light-intensity activities, those that are constituent of ‘everyday’ activities, it may be that there is less of a difference between girls and boys, with limited difference in the deviation or skew.

## Methods

### Sample

The International Children’s Accelerometery Database (ICAD) was used in this analysis.^34^ ICAD is a harmonised dataset of accelerometery data from a series of youth activity studies that employed waist-worn accelerometers in comparable means.^34^ Data was harmonised by reprocessing the raw accelerometer data from each study with a consistent methodology.^35^ Further, social and demographic information were recoded to a consistent reporting, with multiple harmonised variables created for each construct to include as broad of a sample as possible.^35^

This analysis used a subset of the available studies that (a) included individuals aged between 5 and 18, (b) were either cross-sectional or the first wave of a longitudinal study of accelerometery, and (c) were not primarily focussed on an intervention group. The included studies were the Pelotas Birth Cohort (Brazil), National Health and Nutrition Examination Survey (NHANES; USA), the Avon Longitudinal Study of Parents and Children (ALSPAC; UK),^36,37^ European Youth Heart Study (EYHS; Denmark, Estonia, Norway and Portugal), the Kinder-Sportstudie (KISS; Switzerland) and the Healthy Eating and Play Study (HEAPS)/ Children Living in Active Neighbourhoods Project (CLAN; Australia) (Table S 1). In total, our sample includes 15,461 individuals, with 4,615 individuals removed due to incomplete or missing detail (Detail available in Figure S 1). Gender was coded as a binary (boys/girls) variable. Reporting of sex and gender varies across the contributing studies, as such, throughout this study the output is treated as a binary interpretation of gender.

### Measures of Activity

For all individuals, a total count of activity recorded by waist-worn accelerometers was collected, and processed in a consistent manner accounting for differing study protocols.^34^ To standardise for differing wear times, the total counts were converted to a mean number of counts per minute (cpm). To demarcate intensity thresholds, Evenson cut points were used (Sedentary < 101cpm ≤ Light <2296cpm ≤ MVPA). These thresholds have been validated for use in youth samples^38^ and their use enables comparisons with existing research. From this, daily mean volumes of time spent in each threshold were calculated, with MVPA being a sum of time in moderate and vigorous activity. To ensure valid comparisons of wakeful activity participants with a mean recording length greater than 16 hours (960 minutes) of recording per day were removed from analysis of either sedentary or counts per minute. In line with previous research,^39–41^ this was done to remove individuals who wore their device while asleep (Figure S 2, n=1,321; 8.5%).

### Analysis

We conducted descriptive analysis of the distributions of each outcome by gender, and estimated the mean, median, standard deviation and skew. Subsequently, the GAMLSS package in R^32,42^ was used to investigate the relative distribution and variation in volumes of physical activity. We estimated percentage differences in the mean and standard deviation between girls and boys using the normal distribution. To examine differences in the median and skewness, a Box-Cox Cole and Green (BCCG) distribution was used. A Box-Cox transformation transforms skewed data to a normal distribution. The power (λ) required to do so is a measure of the non-normality of the underlying data and is reported here as a measure of skewness. This is estimated by maximum likelihood, and varies by the degree of skew, a power of one is required to transform normal data, with values lower than this for increasingly positive skews, and higher for negative skews.^32^ Given the transformation of skew in a Box-Cox, the measure of central tendency is given by the median. However, due to the log function within BCCG, negative and 0 values cannot be passed, thus any 0 values were recoded to 0.001, this was only necessary for measures of MVPA. GAMLSS requires complete cases to run, as such the sample was restricted to individuals with valid gender, covariate and outcome data.

We first conducted unadjusted models then additionally adjusted for factors which may in part explain or confound gender differences in activity: measured body mass index (kg/m^2^, converted to internally calculated gender and age-specific z-scores^43^), parental education, and study. Heights and weights were taken at the time of accelerometery recording, or from the closest time point to accelerometery. No anthropometrics were taken more than 6 months after accelerometery. Details on both parents’ education was included in all studies except the Pelotas study, for whom only mothers’ education was available. Education was recoded to whether their parents had remained in schooling up to state minimums or whether a parent had received further education, consistent with previous analyses of the ICAD dataset.^21^

To test the robustness of the results, and to ensure this was not a function of non-random missingness, sensitivity analyses were conducted. This involved repeating the unadjusted model with the dataset only restricted to complete cases for activity and gender. Ethnicity (white/other) and season were only available in a subset of individuals (Ethnicity: N= 9,898, 64%; Season: N= 9,604, 62%); we thus adjusted for these factors in additional sensitivity analysis of these countries. MVPA is additionally broken down into moderate and vigorous intensity activities and analysed separately.

## Results

After data cleaning 15,461 (Boys: 48.3%) individuals had complete data for analysis (Table 1). The percentage of boys in studies varied between 45% (Denmark) and 52% (Brazil). The mean age of the sample was 11 years 8 months, ranged from 4.35 to 18.42 years (Table 1) and was similar for boys and girls (Figure S 3). As study design is mixed, there was variation in the variance between countries, with birth cohort studies having a narrower spread of ages. Boys were taller and heavier than girls in the sample, but the differences were negligible (Δ 2cm & Δ 1kg). Of individuals who reported ethnicity, 70% were white, (70.5% of girls, 69.1% of boys) but this information was missing for Australia and Switzerland. Of those responding, roughly 69% of mothers and fathers had education beyond compulsory level, and this was balanced between girls and boys.

**Table 1:**
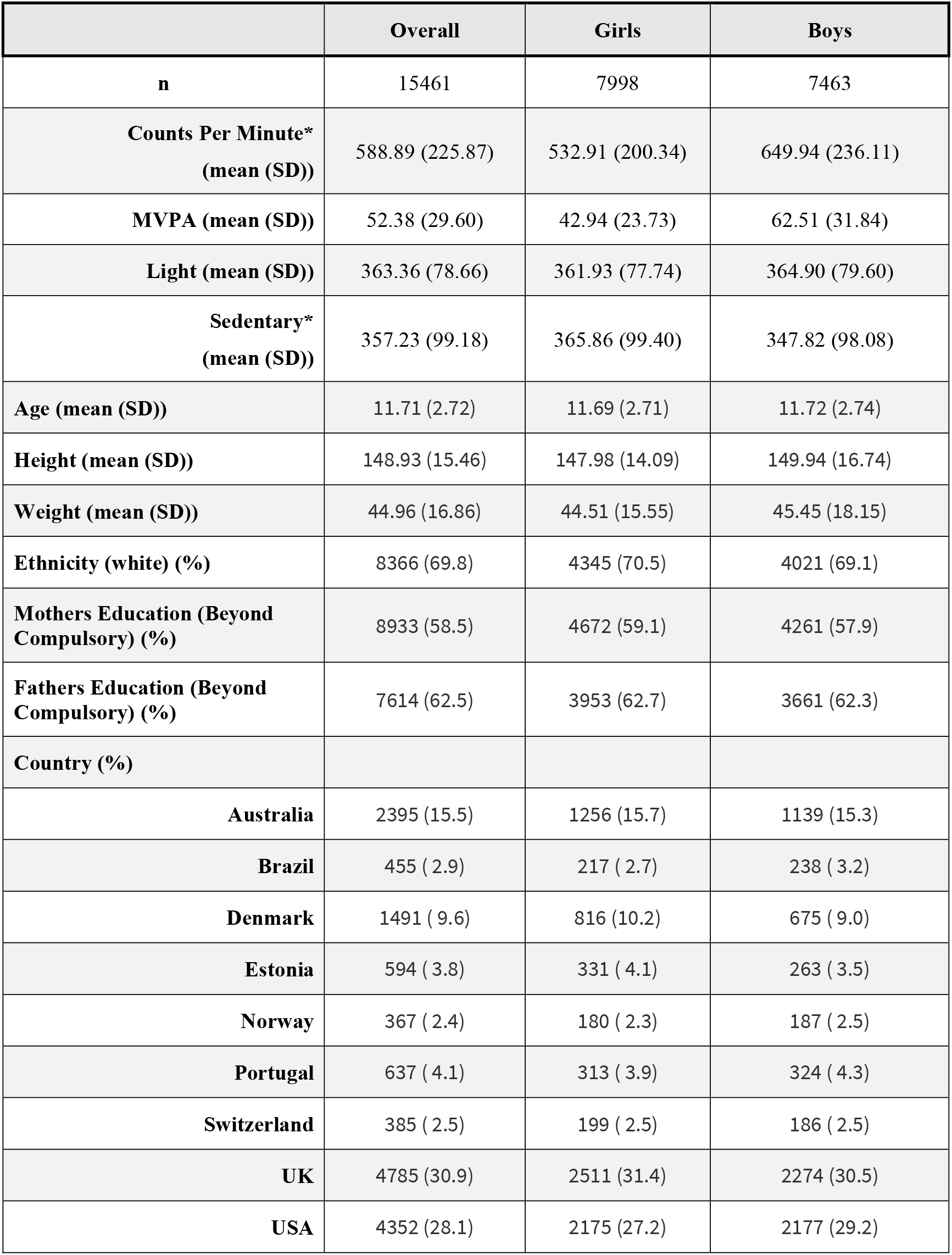
Sample Characteristics, and summary of physical activity volumes stratified by gender. * = Values are reported after exclusion of individuals with more than 16hrs of recording per day. Detail on fathers’ education was absent in the Brazilian (Pelotas) study

|  | Overall | Girls | Boys |
| --- | --- | --- | --- |
| **n** | 15461 | 7998 | 7463 |
| **Counts Per Minute\***<br>**(mean (SD))** | 588.89 (225.87) | 532.91 (200.34) | 649.94 (236.11) |
| **MVPA (mean (SD))** | 52.38 (29.60) | 42.94 (23.73) | 62.51 (31.84) |
| **Light (mean (SD))** | 363.36 (78.66) | 361.93 (77.74) | 364.90 (79.60) |
| **Sedentary\***<br>**(mean (SD))** | 357.23 (99.18) | 365.86 (99.40) | 347.82 (98.08) |
| **Age (mean (SD))** | 11.71 (2.72) | 11.69 (2.71) | 11.72 (2.74) |
| **Height (mean (SD))** | 148.93 (15.46) | 147.98 (14.09) | 149.94 (16.74) |
| **Weight (mean (SD))** | 44.96 (16.86) | 44.51 (15.55) | 45.45 (18.15) |
| **Ethnicity (white) (%)** | 8366 (69.8) | 4345 (70.5) | 4021 (69.1) |
| **Mothers Education (Beyond Compulsory) (%)** | 8933 (58.5) | 4672 (59.1) | 4261 (57.9) |
| **Fathers Education (Beyond Compulsory) (%)** | 7614 (62.5) | 3953 (62.7) | 3661 (62.3) |
| **Country (%)** |  |  |  |
| **Australia** | 2395 (15.5) | 1256 (15.7) | 1139 (15.3) |
| **Brazil** | 455 ( 2.9) | 217 ( 2.7) | 238 ( 3.2) |
| **Denmark** | 1491 ( 9.6) | 816 (10.2) | 675 ( 9.0) |
| **Estonia** | 594 ( 3.8) | 331 ( 4.1) | 263 ( 3.5) |
| **Norway** | 367 ( 2.4) | 180 ( 2.3) | 187 ( 2.5) |
| **Portugal** | 637 ( 4.1) | 313 ( 3.9) | 324 ( 4.3) |
| **Switzerland** | 385 ( 2.5) | 199 ( 2.5) | 186 ( 2.5) |
| **UK** | 4785 (30.9) | 2511 (31.4) | 2274 (30.5) |
| **USA** | 4352 (28.1) | 2175 (27.2) | 2177 (29.2) |

### Counts Per Minute

Boys recorded over 100cpm more than girls on average (Girls: 532.91cpm, Boys: 649.94cpm; Table 1), with estimated values that were 20% greater than that of girls for both the mean and median in the adjusted model (Table 2). Boys had a wider distribution of values (Figure 1 A), marked by a greater standard deviation (Girls: 200.34, Boys: 236.11); 17% larger (Table 2). Both girls and boys had moderately positive measures of skew, with the strength of skew marginally stronger for girls (as noted by a score closer to 1 in Table 2). The results were broadly similar across all 9 countries (Figure S 5), with the exception of Portugal (EYHS) for which little inequality in the distribution was observed between girls and boys.

**Table 2:** Association between gender and physical activity as measured by counts per minute. Differences in mean, variability and skew estimated by GAMLSS, a = adjusted for parental education, BMI, and country. NO: normal distribution. b = Skewness is estimated as the Box-Cox power (that is, the power required to transform the outcome to a normal distribution, values closer to 1 represent less skew). BCCG: Box-Cox Cole and Green distribution: SD: standard deviation. GAMLSS: Generalized Additive Models for Location, Scale and Shape. SE, standard error. * = p<0.05, ** = p<0.01, *** = p<0.001.

| Gender | N (%) | NO distribution |  | BCCG distribution |  |
| --- | --- | --- | --- | --- | --- |
|  |  | Mean<br>% Difference<br>(SE) | SD<br>% Difference<br>(SE) | Median<br>% Difference<br>(SE) | Skewness<sup>b</sup> |
| **Girls (ref)**<br>Unadjusted | 7377<br>(52.2) | 532.91 | 200.32 | 513.31 | 0.48 |
| **Boys**<br>(Unadjusted Difference) | 6763<br>(47.8) | 19.85 (0.62)<br>*** | 16.42 (1.26)<br>*** | 21.29 (0.67)<br>*** | 0.19 (0.03)<br>*** |
| **Boys<sup>a</sup>**<br>(Adjusted Difference) | 6763<br>(47.8) | 19.74 (0.58)<br>*** | 17.24 (1.19)<br>*** | 21.14 (0.63)<br>*** | 0.17 (0.03)<br>*** |

**Figure 1:**
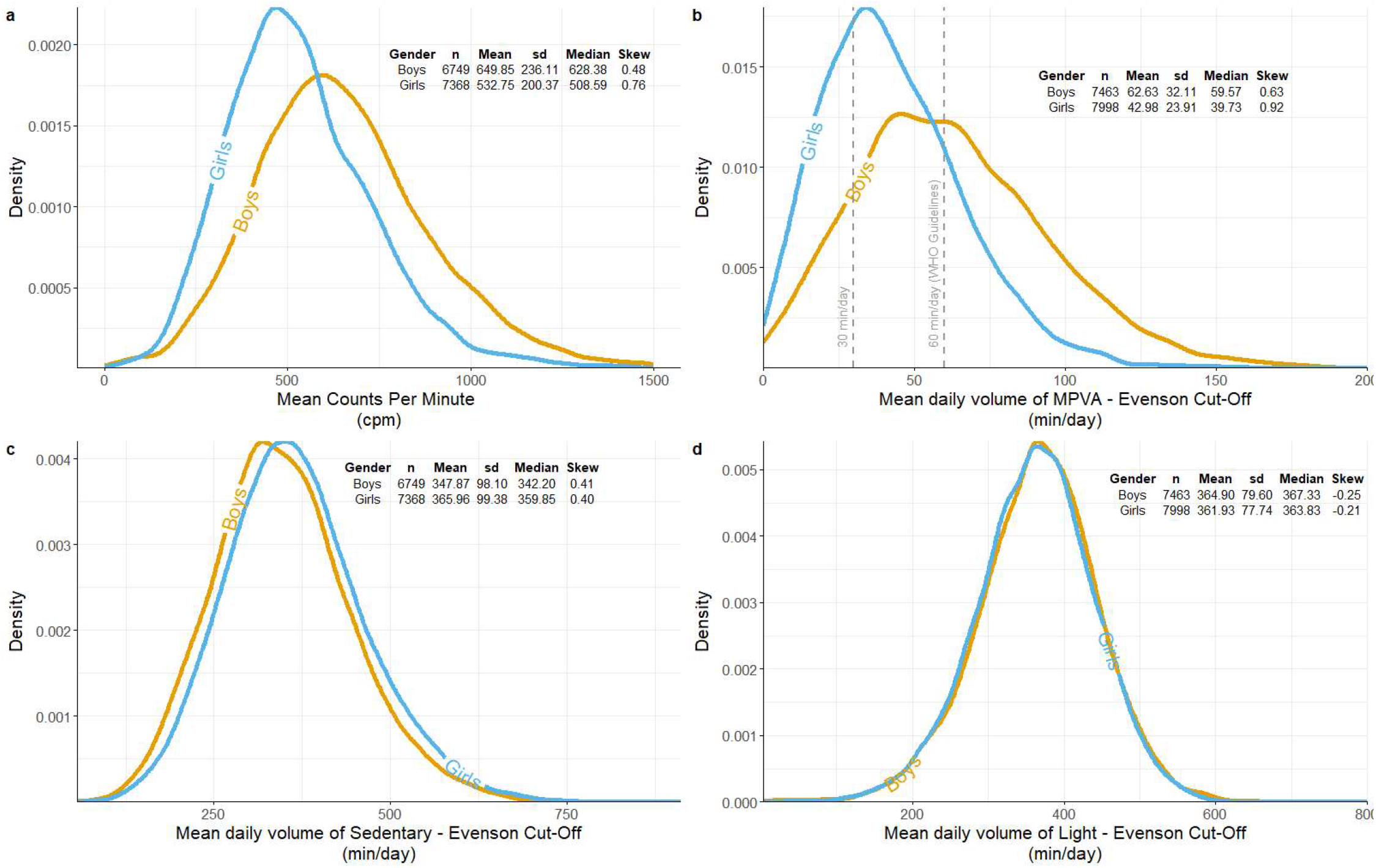
Density plot of each activity measure with each line representing a gender. (A) displays mean counts per minute by gender, plot is muted at 1,500 cpm to centre of distribution. (B) displays moderate-to-vigorous activity as defined by Evenson cut points. Plot is muted at 200 min/day to show centre of distribution. 60 minutes per day and 30 minutes per day are marked by vertical dashed lines. (C) displays light-intensity activity as defined by Evenson cut points. (D) displays sedentary activity as defined by Evenson cut points.

### Moderate to Vigorous Physical Activity (MVPA)

Using Evenson cut points,^38^ boys recorded a greater mean (Girls: 42.94 min/day, Boys: 62.51 min/day; Table 1) and median (Girls: 39.71 min/day, Boys: 59.50 min/day) volume of MVPA (Figure 1 B). By this measure the mean boy was meeting health guidelines of 60 minutes of MVPA per day, with the mean girl reporting 20 minutes less than the same target per day. In the adjusted model, boys recorded a mean volume 37% greater and a median 40% greater than girls (Table 3). Boys showed more variation in their daily volumes of MPVA with a greater standard deviation (Girls: 23.73, Boys: 31.84; Figure 1 B), with an estimated value 30% greater for boys in the adjusted model (Table 3). Both distributions were positively skewed, with the strength of skew slightly stronger for girls than boys (Girls: 0.82, Boys: 0.57; Figure 1 B). As with counts per minute, the results were broadly comparable on a country-by-country level.

**Table 3:**
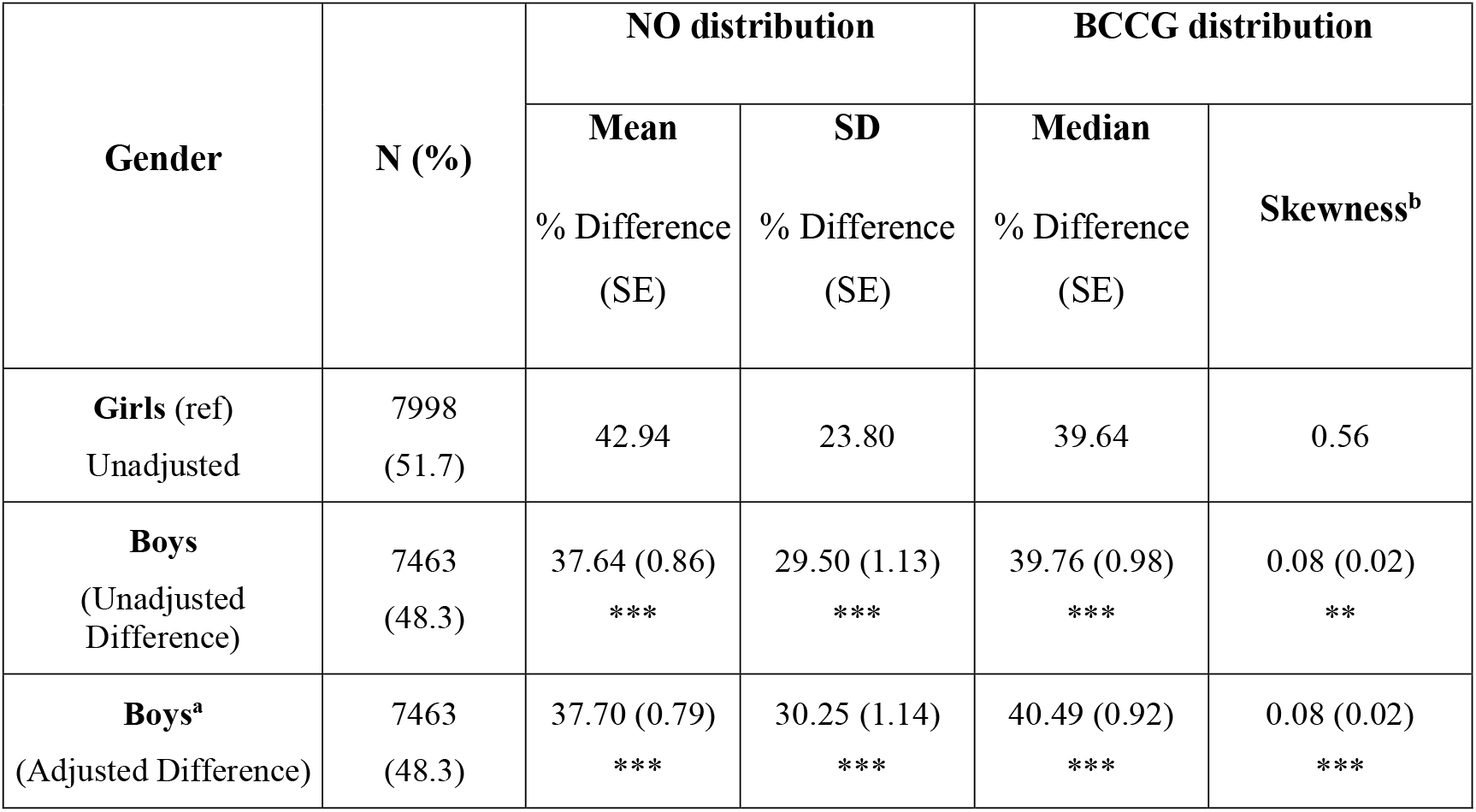
Association between gender and moderate to vigorous activity as defined by Evenson cut points. Differences in mean, variability and skew estimated by GAMLSS, a = adjusted for parental education, BMI, and country. NO: normal distribution. b = Skewness is estimated as the Box-Cox power (that is, the power required to transform the outcome to a normal distribution, values closer to 1 represent less skew). BCCG: Box-Cox Cole and Green distribution: SD: standard deviation. GAMLSS: Generalized Additive Models for Location, Scale and Shape. SE, standard error. * = p<0.05, ** = p<0.01, *** = p<0.001.

| Gender | N (%) | NO distribution |  | BCCG distribution |  |
| --- | --- | --- | --- | --- | --- |
|  |  | Mean<br>% Difference<br>(SE) | SD<br>% Difference<br>(SE) | Median<br>% Difference<br>(SE) | Skewness<sup>b</sup> |
| **Girls (ref)**<br>Unadjusted | 7998<br>(51.7) | 42.94 | 23.80 | 39.64 | 0.56 |
| **Boys**<br>(Unadjusted<br>Difference) | 7463<br>(48.3) | 37.64 (0.86)<br>*** | 29.50 (1.13)<br>*** | 39.76 (0.98)<br>*** | 0.08 (0.02)<br>** |
| **Boys<sup>a</sup>**<br>(Adjusted Difference) | 7463<br>(48.3) | 37.70 (0.79)<br>*** | 30.25 (1.14)<br>*** | 40.49 (0.92)<br>*** | 0.08 (0.02)<br>*** |

### Sedentary and Light-intensity Activity

Girls and boys had similar distributions for sedentary and light-intensity activities; both undertook approximately 6hrs of sedentary (Girls: 368.74min/day, Boys: 350.17min/day) and light-intensity activity per day (Girls: 358.58 min/day, Boys: 364.90 min/day; Figure 1 C & D). The differences were marginal with girls estimated to have 5% higher volumes of sedentary activity, and less than 1% lower in light-intensity activity than boys (Table 4 & 5). Standard deviations were closely aligned for both sedentary (Girls: 98.79, Boys: 97.92) and light-intensity activity (Girls: 77.74, Boys: 79.60) with an estimated difference of 1% and 3% respectively in the adjusted models (Table 4 & 5**Error! Reference source not found**.). Data for both girls and boys were relatively normally distributed, being weakly positively skewed for sedentary behaviour (Table 4) with low, negative measures of skew for light-intensity activity (**Error! Reference source not found**.Table 5). Within each individual nation, the observed patterns of light activity for girls and boys were broadly similar (Figure S 8).

**Table 4:**
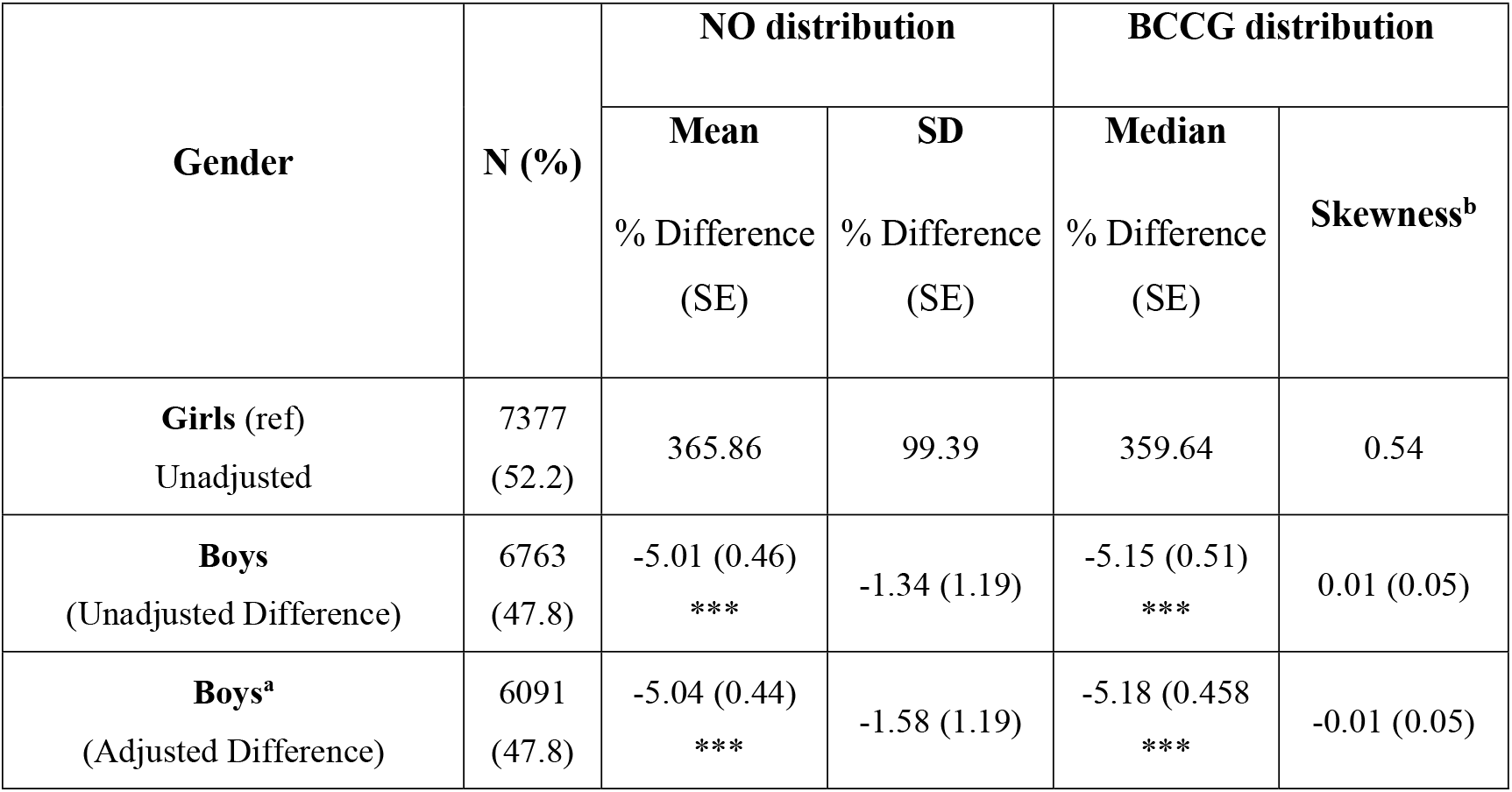
Association between gender and sedentary activity as defined by Evenson cut points. Differences in mean, variability and skew estimated by GAMLSS, a = adjusted for parental education, BMI, and country. NO: normal distribution. b = Skewness is estimated as the Box-Cox power (that is, the power required to transform the outcome to a normal distribution, values closer to 1 represent less skew). BCCG: Box-Cox Cole and Green distribution: SD: standard deviation. CoV: coefficient of variation. GAMLSS: Generalized Additive Models for Location, Scale and Shape. SE, standard error. * = p<0.05, ** = p<0.01,*** = p<0.001.

**Table 5:** Association between gender and light-intensity activity as defined by Evenson cut points. Differences in mean, variability and skew estimated by GAMLSS, a = adjusted for parental education, BMI, and country. NO: normal distribution. b = Skewness is estimated as the Box-Cox power (that is, the power required to transform the outcome to a normal distribution, values closer to 1 represent less skew). BCCG: Box-Cox Cole and Green distribution: SD: standard deviation. GAMLSS: Generalized Additive Models for Location, Scale and Shape. SE, standard error. * = p<0.05, ** = p<0.01, *** = p<0.001.

| Gender | N (%) | NO distribution |  | BCCG distribution |  |
| --- | --- | --- | --- | --- | --- |
|  |  | Mean<br>% Difference<br>(SE) | SD<br>% Difference<br>(SE) | Median<br>% Difference<br>(SE) | Skewness<sup>b</sup> |
| **Girls (ref)**<br>Unadjusted | 7998<br>(51.7) | 361.93 | 77.73 | 364.43 | 1.29 |
| **Boys**<br>(Unadjusted Difference) | 7463 (48.3) | 0.82 (0.34)<br>* | 2.36 (1.14)<br>* | 0.90 (0.36)<br>* | 0.03 (0.05) |
| **Boys<sup>a</sup>**<br>(Adjusted Difference) | 7463 (48.3) | 0.88 (0.33)<br>** | 3.26 (1.14)<br>** | 0.87 (0.35)<br>* | -0.02 (0.06) |

### Sensitivity Analysis

Results were similar when repeating the unadjusted model with the unrestricted dataset (n=18,980; Table S 3-6). Adjustment for season and ethnicity made little difference, leading to similar estimated effect sizes of gender for any measure (Table S 7-10). Separating MVPA into moderate and vigorous intensity activity resulted in similar effects, that were larger for vigorous activities (Table S 11 & 12).

## Discussion

### Summary of Findings

Boys recorded greater mean activity levels than girls but with higher variability, revealing more inequality in activity within boys than girls. This was driven by differences in MVPA: boys spent more time on average in MVPA, with greater variation between them. More equality amongst girls at a lower mean volume implied that few girls in this sample were doing large amounts of MVPA, resulting in a narrow spread of girls centred around median volumes of non-volitional of MVPA.

In contrast, for both sedentary and light-intensity activity, the differences in the distribution for girls and boys were marginal, with little inequality in the variation for either measure or for the total volumes. These intensities form most of the wakeful time, indicating that differences in overall activity (cpm) are driven by a small subset of daily behaviours in moderate to vigorous thresholds.

### Explanation of Findings

Consistent with previous studies, girls and boys differed in their mean activity count and volume of MVPA.^18–24^ However, the lack of difference in the volume of light-intensity activity implies that the difference between girls and boys in their overall activity was unlikely to be due to ‘every day’ activities that characterises the light-intensity spectrum, but instead are driven by changes at the upper end of the spectrum, which in context of the included populations would likely align with sports and active play.^44–46^

Adding to the existing body of research is a quantification of the difference in the distribution. We observed a higher mean volume of MVPA for boys alongside a higher standard deviation suggesting that not only was the average boy more active than the average girl, more boys occupied the highest volumes of activity (Figure 1 B). Volumes of MVPA were more homogenous in girls, but clustered around a lower value. A possible explanation of this is that there was less inequality in volumes of ‘day-to-day’ MVPA (i.e. active commutes or physical education classes; In Figure 1 B the volume of MVPA at which the peak density is observed is relatively close between girls and boys), but a larger proportion of boys were undertaking additional volitional activity, increasing the inequality between genders and within boys.^47^

As such, when interpreting differences between girls and boys in mean volumes of activity, changes were not driven by the whole population, but instead driven by the subset undertaking additional activity. The larger subset of boys undertaking near daily volitional sports or active play for at least an hour per day on top of their other activities provided more balance to the distribution despite the increased heterogeneity, resulting in a larger standard deviation yet lower skew.

While almost all countries show similar gender differences, Portugal did not have as much of a gender divide in total activity (cpm) as other nations did (Figure S 2). Within this sample boys were less active when compared to other constituent studies than girls, with Portuguese boys ranked 8th (of 8) in median counts per minute compared to 5th for Portuguese girls. While girls and boys may have equal opportunity to undertake non-volitional activity, limited access to sport and leisure facilities on the island of Madeira^48^ may equalise opportunities for individuals to engage in volitional activity,^49^ reducing the opportunities for a difference between boys and girls to emerge. Alternatively, it may be that on Madeira, the volitional activities that would differentiate individuals are not accounted for by the mode of accelerometery, such as swimming or cycling.

### Potential Implications

Deficits between girls and boys in total activity observed here (measured by CPM) were derived from difference in MVPA. If the deficit was due to a lack of opportunities for sport and active play for girls, targeting interventions at higher intensity activities could be central to reducing the inequality between girls and boys. However, such interventions could increase opportunities for those who are already somewhat active but lead to little change amongst the least active. If that is the case, then increasing opportunities for volitional activity for girls may inadvertently act to increase the inequality within girls. To maximise the efficacy of sport-based interventions it may also be necessary to address the socio-cultural reasons for lower uptake of active play and sport by girls.^33,50^

Beyond differences between girls, activity remains low for most children both in this study and in other research.^16,17^ Given the similarity between girls and boys in light-intensity activity (Figure 1 D) repeated across all constituent studies in the present research (Figure S 5), it may be that an intervention that targets light-intensity activity, such as improving community walkability, is more likely to have an impact for both genders than an intervention that targets more strenuous activity (such as sports).

### Strengths and Limitations

The sample size in this study was extensive, spanning multiple countries, which suggests that results may be generalisable across multiple cultures. Further, using harmonised, objective measures of physical activity captured a large range of activity at multiple intensities, rather than behaviours that are challenging to accurately recall. Processing this data in a consistent manner avoided issues associated with comparing between differing methodologies. Finally, by employing a GAMLSS analysis across all intensity thresholds, differences between the distribution were tested, allowing difference in the spread and skew of samples to be directly examined.

GAMLSS has limitations to its use; the requirement of complete cases restricts the sample size throughout. While this presents a risk that the data was not missing at random, no notable changes in the results were seen during sensitivity analyses. Accelerometery is limited in its ability to measure resistance-based activities, and activities like cycling. Of the included studies, this may have led to underestimated volumes of activity in Danish EYHS study, in which most students cycle to school multiple times per week.^51^ However rates of cycling are not expected to differ between boys and girls.^52^ Further, to ensure backward compatibility between different accelerometers, recordings from newer devices were re-processed at the resolution of the oldest devices coming at the cost of some resolution.^34^ As the sample draws from countries with broadly similar political, economic and cultural practices, further work is needed in low-to-middle income nations.^53^

## Conclusions

In addition to inequality between girls and boys, we observed sizable and variable levels of inequality within each gender. Differences in overall activity (cpm) were mainly driven by the upper end of the activity spectrum; both the higher intensity activities (MVPA) and the most active individuals. Boys were more active on average, but this is due to a sizable subset of boys undertaking high volumes of MVPA, rather than all boys doing a small amount more than girls. Attention should therefore be placed on the full distribution of individuals, as an intervention may narrow the difference between boys and girls yet, by focussing this change on a subset of individuals, it may exacerbate the inequality within boys and girls. For equitable change in children’s activity, interventions should aim to benefit those in the lowest quantiles as effectively as the highest.

## Data Availability

All data produced are available online at https://www.mrc-epid.cam.ac.uk/research/studies/icad/

https://rpubs.com/LukeKretschmer/1027505

## Acknowledgements

We would like to thank all participants and funders of the original studies that contributed data to ICAD. We gratefully acknowledge the past contributions of Prof Chris Riddoch, Prof Ken Judge, Prof Ashley Cooper and Dr Pippa Griew to the development of ICAD.

The ICAD was made possible thanks to the sharing of data from the following contributors (study name): Prof LB Andersen, Faculty of Teacher Education and Sport, Western Norway University of Applied Sciences, Sogndal, Norway (Copenhagen School Child Intervention Study (CoSCIS)); Prof S Anderssen, Norwegian School for Sport Science, Oslo, Norway (European Youth Heart Study (EYHS), Norway); Prof G Cardon, Department of Movement and Sports Sciences, Ghent University, Belgium (Belgium Pre-School Study); Centers for Disease Control and Prevention (CDC), National Center for Health Statistics (NCHS), Hyattsville, MD USA (National Health and Nutrition Examination Survey (NHANES)); Dr R Davey, Centre for Research and Action in Public Health, University of Canberra, Australia (Children’s Health and Activity Monitoring for Schools (CHAMPS)); Prof. Pedro C Hallal, Department of Kinesiology and Community Health, University of Illinois Urbana-Champaign, United States (1993 Pelotas Birth Cohort); Prof R Jago, Centre for Exercise, Nutrition and Health Sciences, University of Bristol, UK (Personal and Environmental Associations with Children’s Health (PEACH)); Prof KF Janz, Department of Health and Human Physiology, Department of Epidemiology, University of Iowa, Iowa City, US (Iowa Bone Development Study); Prof S Kriemler, Epidemiology, Biostatistics and Prevention Institute, University of Zürich, Switzerland (Kinder-Sportstudie (KISS)); Dr N Møller, University of Southern Denmark, Odense, Denmark (European Youth Heart Study (EYHS), Denmark); Prof K Northstone, School of Social and Community Medicine, University of Bristol, UK (Avon Longitudinal Study of Parents and Children (ALSPAC)); Prof R Pate, Department of Exercise Science, University of South Carolina, Columbia, US (Physical Activity in Pre-school Children (CHAMPS-US) and Project Trial of Activity for Adolescent Girls (Project TAAG)); Dr JJ Puder, Service of Endocrinology, Diabetes and Metabolism, Centre Hospitalier Universitaire Vaudois, University of Lausanne, Switzerland (Ballabeina Study); Prof J Reilly, Physical Activity for Health Group, School of Psychological Sciences and Health, University of Strathclyde, Glasgow, UK (Movement and Activity Glasgow Intervention in Children (MAGIC)); Prof J Salmon, Institute for Physical Activity and Nutrition (IPAN), School of Exercise and Nutrition Sciences, Deakin University, Geelong, Australia (Children Living in Active Neigbourhoods (CLAN) & Healthy Eating and Play Study (HEAPS)); Prof LB Sardinha, Exercise and Health Laboratory, Faculty of Human Movement, Universidade de Lisboa, Lisbon, Portugal (European Youth Heart Study (EYHS), Portugal); Dr EMF van Sluijs, MRC Epidemiology Unit & Centre for Diet and Activity Research, University of Cambridge, UK (Sport, Physical activity and Eating behaviour: Environmental Determinants in Young people (SPEEDY)). Thanks are also extended to Evangeline Tabor (UCL, UK) for her advice on the manuscript

The pooling of the data was funded through a grant from the National Prevention Research Initiative (Grant Number: G0701877) (http://www.mrc.ac.uk/research/initiatives/national-prevention-research-initiative-npri/). The funding partners relevant to this award are: British Heart Foundation; Cancer Research UK; Department of Health; Diabetes UK; Economic and Social Research Council; Medical Research Council; Research and Development Office for the Northern Ireland Health and Social Services; Chief Scientist Office; Scottish Executive Health Department; The Stroke Association; Welsh Assembly Government and World Cancer Research Fund. This work was additionally supported by the Medical Research Council [MC_UU_12015/3; MC_UU_12015/7], The Research Council of Norway (249932/F20), Bristol University, Loughborough University and Norwegian School of Sport Sciences.

LK is funded by the ESRC-BBSRC Soc-B Centre for Doctoral Training (ES/P000347/1)

DB is supported by the Economic and Social Research Council (grant number ES/M001660/1) and Medical Research Council (MR/V002147/1).

## References

1. World Health Organization. (2020). WHO Guidelines On Physical Activity And Sedentary Behaviour.

2. Timmons, B. W. (2016). Exercise and Immune Function in Children: http://Dx.Doi.Org/10.1177/1559827606294851, 1(1), p59–66.

3. Janssen, I., & LeBlanc, A. G. (2010). Systematic review of the health benefits of physical activity and fitness in school-aged children and youth. International Journal of Behavioral Nutrition and Physical Activity, 7(1), 1–16.

4. Owen, C. G., Nightingale, C. M., Rudnicka, A. R., Sattar, N., Cook, D. G., Ekelund, U., & Whincup, P. H. (2010). Physical activity, obesity and cardiometabolic risk factors in 9-to 10-year-old UK children of white European, South Asian and black African-Caribbean origin: The Child Heart and health Study in England (CHASE). Diabetologia, 53(8), 1620–1630.

5. Platat, C., Wagner, A., Klumpp, T., Schweitzer, B., & Simon, C. (2006). Relationships of physical activity with metabolic syndrome features and low-grade inflammation in adolescents. Diabetologia, 49(9), 2078–2085.

6. Ruiz, J. R., Ortega, F. B., Warnberg, J., & Sjöström, M. (2007). Associations of low-grade inflammation with physical activity, fitness and fatness in prepubertal children; The European Youth Heart Study. International Journal of Obesity, 31(10), 1545–1551.

7. Poitras, V. J., Gray, C. E., Borghese, M. M., Carson, V., Chaput, J. P., Janssen, I., Katzmarzyk, P. T., Pate, R. R., Connor Gorber, S., Kho, M. E., Sampson, M., & Tremblay, M. S. (2016). Systematic review of the relationships between objectively measured physical activity and health indicators in school-aged children and youth. Applied Physiology, Nutrition and Metabolism, 41(6), S197–S239.

8. García-Hermoso, A., Ezzatvar, Y., Ramírez-Vélez, R., Olloquequi, J., & Izquierdo, M. (2020). Is device-measured vigorous-intensity physical activity associated with health-related outcomes in children and adolescents? A systematic review and meta-analysis. Journal of Sport and Health Science.

9. Biddle, S. J. H., & Asare, M. (2011). Physical activity and mental health in children and adolescents: A review of reviews. British Journal of Sports Medicine, 45(11), 886–895.

10. Veldman, S. L. C., Chin A Paw, M. J. M., & Altenburg, T. M. (2021). Physical activity and prospective associations with indicators of health and development in children aged <5 years: a systematic review. International Journal of Behavioral Nutrition and Physical Activity, 18(1), 1–11.

11. Gunter, K. B., Almstedt, H. C., & Janz, K. F. (2012). Physical activity in childhood may be the key to optimizing lifespan skeletal health. Exercise and Sport Sciences Reviews, 40(1), 13–21.

12. Carter, M. I., & Hinton, P. S. (2014). Physical activity and bone health. Missouri Medicine, 111(1), 59–64.

13. Bass, S., Pearce, G., Bradney, M., Hendrich, E., Delmas, P. D., Harding, A., & Seeman, E. (1998). Exercise Before Puberty May Confer Residual Benefits in Bone Density in Adulthood: Studies in Active Prepubertal and Retired Female Gymnasts. Journal of Bone and Mineral Research, 13(3), 500–507.

14. Bland, V. L., Heatherington-Rauth, M., Howe, C., Going, S. B., & Bea, J. W. (2020). Association of objectively measured physical activity and bone health in children and adolescents: a systematic review and narrative synthesis. Osteoporosis International, 1–30.

15. Leão, O. A. de A., Mielke, G. I., Hallal, P. C., Cairney, J., Mota, J., Domingues, M. R., Murray, J., & Bertoldi, A. D. (2022). Longitudinal Associations Between Device-Measured Physical Activity and Early Childhood Neurodevelopment. Journal of Physical Activity and Health, 1(aop), 1–9.

16. Bauman, A. E., Reis, R. S., Sallis, J. F., Wells, J. C., Loos, R. J. F., Martin, B. W., Alkandari, J. R., Andersen, L. B., Blair, S. N., Brownson, R. C., Bull, F. C., Craig, C. L., Ekelund, U., Goenka, S., Guthold, R., Hallal, P. C., Haskell, W. L., Heath, G. W., Inoue, S., … Sarmiento, O. L. (2012). Correlates of physical activity: Why are some people physically active and others not? The Lancet, 380(9838), 258–271.

17. Trost, S. G., Pate, R. R., Sallis, J. F., Freedson, P. S., Taylor, W. C., Dowda, M., & Sirard, J. (2002). Age and gender differences in objectively measured physical activity in youth. In Med. Sci. Sports Exerc (Vol. 34, Issue 2, pp. 350–355). http://www.acsm-msse.org

18. Cooper, A. R., Goodman, A., Page, A. S., Sherar, L. B., Esliger, D. W., van Sluijs, E. M. F., Andersen, L. B., Anderssen, S., Cardon, G., Davey, R., Froberg, K., Hallal, P., Janz, K. F., Kordas, K., Kreimler, S., Pate, R. R., Puder, J. J., Reilly, J. J., Salmon, J., … Ekelund, U. (2015). Objectively measured physical activity and sedentary time in youth: The International children’s accelerometry database (ICAD). International Journal of Behavioral Nutrition and Physical Activity, 12(1), 1–10.

19. Corder, K., Sharp, S. J., Atkin, A. J., Andersen, L. B., Cardon, G., Page, A., Davey, R., Grøntved, A., Hallal, P. C., Janz, K. F., Kordas, K., Kriemler, S., Puder, J. J., Sardinha, L. B., Ekelund, U., van Sluijs, E. M. F., Cardon, G., Cooper, A., Puder, J. J., … Timperio, A. (2016). Age-related patterns of vigorous-intensity physical activity in youth: The International Children’s Accelerometry Database. Preventive Medicine Reports, 4, 17–22.

20. Steene-Johannessen, J., Hansen, B. H., Dalene, K. E., Kolle, E., Northstone, K., Møller, N. C., Grøntved, A., Wedderkopp, N., Kriemler, S., Page, A. S., Puder, J. J., Reilly, J. J., Sardinha, L. B., Van Sluijs, E. M. F., Andersen, L. B., Van Der Ploeg, H., Ahrens, W., Flexeder, C., Standl, M., … Van Sluijs, E. M. F. (2020). Variations in accelerometry measured physical activity and sedentary time across Europe-harmonized analyses of 47,497 children and adolescents. International Journal of Behavioral Nutrition and Physical Activity, 17(1), 1–14.

21. Dias, K. I., White, J., Jago, R., Cardon, G., Davey, R., Janz, K. F., Pate, R. R., Puder, J. J., Reilly, J. J., & Kipping, R. (2019). International Comparison of the Levels and Potential Correlates of Objectively Measured Sedentary Time and Physical Activity among Three-to-Four-Year-Old Children. International Journal of Environmental Research and Public Health 2019, Vol. 16, Page 1929, 16(11), 1929.

22. Van Ekris, E., Wijndaele, K., Altenburg, T. M., Atkin, A. J., Twisk, J., Andersen, L. B., Janz, K. F., Froberg, K., Northstone, K., Page, A. S., Sardinha, L. B., Van Sluijs, E. M. F., Chinapaw, M., Andersen, L. B., Anderssen, S., Atkin, A. J., Cardon, G., Davey, R., Ekelund, U., … Van Sluijs, E. M. F. (2020). Tracking of total sedentary time and sedentary patterns in youth: A pooled analysis using the International Children’s Accelerometry Database (ICAD). International Journal of Behavioral Nutrition and Physical Activity, 17(1), 1–10.

23. Kwon, S., Janz, K. F., Cooper, A., Ekelund, U., Esliger, D., Griew, P., Judge, K., Ness, A., Riddoch, C., Salmon, J., & Sherar, L. (2012). Tracking of accelerometry-measured physical activity during childhood: ICAD pooled analysis. International Journal of Behavioral Nutrition and Physical Activity, 9(1), 1–8.

24. Tarp, J., Child, A., White, T., Westgate, K., Bugge, A., Grøntved, A., Wedderkopp, N., Andersen, L. B., Cardon, G., Davey, R., Janz, K. F., Kriemler, S., Northstone, K., Page, A. S., Puder, J. J., Reilly, J. J., Sardinha, L. B., van Sluijs, E. M. F., Ekelund, U., … Brage, S. (2018). Physical activity intensity, bout-duration, and cardiometabolic risk markers in children and adolescents. International Journal of Obesity 2018 42:9, 42(9), 1639–1650.

25. Konstabel, K., Veidebaum, T., Verbestel, V., Moreno, L. A., Bammann, K., Tornaritis, M., Eiben, G., Molnár, D., Siani, A., Sprengeler, O., Wirsik, N., Ahrens, W., & Pitsiladis, Y. (2014). Objectively measured physical activity in European children: the IDEFICS study. International Journal of Obesity 2014 38:2, 38(2), S135–S143.

26. Hallal, P. C., Victora, C. G., Azevedo, M. R., & Wells, J. C. K. (2006). Adolescent physical activity and health: A systematic review. Sports Medicine, 36(12), 1019–1030.

27. Guthold, R., Stevens, G. A., Riley, L. M., & Bull, F. C. (2018). Worldwide trends in insufficient physical activity from 2001 to 2016: a pooled analysis of 358 population-based surveys with 1·9 million participants. The Lancet Global Health, 6(10), e1077–e1086.

28. Chaput, J.-P., Barnes, J. D., Tremblay, M. S., Fogelholm, M., Hu, G., Lambert, E. V., Maher, C., Maia, J., Olds, T., Onywera, V., Sarmiento, O. L., Standage, M., Tudor-Locke, C., & Katzmarzyk, P. T. (2018). Inequality in physical activity, sedentary behaviour, sleep duration and risk of obesity in children: a 12-country study. Obesity Science & Practice, 4(3), 229–237.

29. Steene-Johannessen, J., Anderssen, S. A., Kolle, E., Hansen, B. H., Bratteteig, M., Dalhaug, E. M., Andersen, L. B., Nystad, W., Ekelund, U., & Dalene, K. E. (2021). Temporal trends in physical activity levels across more than a decade - a national physical activity surveillance system among Norwegian children and adolescents. The International Journal of Behavioral Nutrition and Physical Activity, 18(1), 55.

30. Basterfield, L., Adamson, A. J., Frary, J. K., Parkinson, K. N., Pearce, M. S., & Reilly, J. J. (2011). Longitudinal Study of Physical Activity and Sedentary Behavior in Children. Pediatrics, 127(1), e24–e30.

31. Bann, D., Wright, L., & Cole, T. J. (2022). Risk factors relate to the variability of health outcomes as well as the mean: A GAMLSS tutorial. ELife, 11.

32. Rigby, R. A., Stasinopoulos, D. M., & Lane, P. W. (2005). Generalized additive models for location, scale and shape. Journal of the Royal Statistical Society: Series C (Applied Statistics), 54(3), 507–554.

33. Chalabaev, A., Sarrazin, P., Fontayne, P., Boiché, J., & Clément-Guillotin, C. (2012). The influence of sex stereotypes and gender roles on participation and performance in sport and exercise: Review and future directions.

34. Sherar, L. B., Griew, P., Esliger, D. W., Cooper, A. R., Ekelund, U., Judge, K., & Riddoch, C. (2011). International children’s accelerometry database (ICAD): Design and methods. BMC Public Health, 11(1), 1–13.

35. Atkin, A. J., Biddle, S. J. H., Broyles, S. T., Chinapaw, M., Ekelund, U., Esliger, D. W., Hansen, B. H., Kriemler, S., Puder, J. J., Sherar, L. B., van Sluijs, E. M. F., Andersen, L. B., Anderssen, S., Cardon, G., Davey, R., Hallal, P., Janz, K. F., Møller, N., Molloy, L., … Timperio, A. (2017). Harmonising data on the correlates of physical activity and sedentary behaviour in young people: Methods and lessons learnt from the international Children’s Accelerometry database (ICAD). International Journal of Behavioral Nutrition and Physical Activity, 14(1), 1–12.

36. Fraser, A., Macdonald-Wallis, C., Tilling, K., Boyd, A., Golding, J., Davey Smith, G., Henderson, J., Macleod, J., Molloy, L., Ness, A., Ring, S., Nelson, S. M., & Lawlor, D. A. (2013). Cohort Profile: The Avon Longitudinal Study of Parents and Children: ALSPAC mothers cohort. International Journal of Epidemiology, 42(1), 97–110.

37. Boyd, A., Golding, J., Macleod, J., Lawlor, D. A., Fraser, A., Henderson, J., Molloy, L., Ness, A., Ring, S., & Davey Smith, G. (2013). Cohort Profile: The ‘Children of the 90s’—the index offspring of the Avon Longitudinal Study of Parents and Children. International Journal of Epidemiology, 42(1), 111–127.

38. Evenson, K. R., Catellier, D. J., Gill, K., Ondrak, K. S., & McMurray, R. G. (2008). Calibration of two objective measures of physical activity for children. Journal of Sports Sciences, 26(14), 1557–1565.

39. Hildebrand, M., Kolle, E., Hansen, B. H., Collings, P. J., Wijndaele, K., Kordas, K., Cooper, A. R., Sherar, L. B., Andersen, L. B., Sardinha, L. B., Kriemler, S., Hallal, P., Van Sluijs, E., & Ekelund, U. (2015). Association between birth weight and objectively measured sedentary time is mediated by central adiposity: data in 10,793 youth from the International Children’s Accelerometry Database. The American Journal of Clinical Nutrition, 101(5), 983–990.

40. Hansen, B. H., Anderssen, S. A., Andersen, L. B., Hildebrand, M., Kolle, E., Steene-Johannessen, J., Kriemler, S., Page, A. S., Puder, J. J., Reilly, J. J., Sardinha, L. B., van Sluijs, E. M. F., Wedderkopp, N., Ekelund, U., Andersen, L. B., Anderssen, S. A., Atkin, A. J., Davey, R., Esliger, D. W., … Timperio, A. (2018). Cross-Sectional Associations of Reallocating Time Between Sedentary and Active Behaviours on Cardiometabolic Risk Factors in Young People: An International Children’s Accelerometry Database (ICAD) Analysis. Sports Medicine, 48(10), 2401–2412.

41. Aadland, E., Kvalheim, O. M., Hansen, B. H., Kriemler, S., Ried-Larsen, M., Wedderkopp, N., Sardinha, L. B., Møller, N. C., Hallal, P. C., Anderssen, S. A., Northstone, K., & Andersen, L. B. (2020). The multivariate physical activity signature associated with metabolic health in children and youth: An International Children’s Accelerometry Database (ICAD) analysis. Preventive Medicine, 141, 106266.

42. R Core Team. (2020). R: A Language and Environment for Statistical Computing. R Foundation for Statistical Computing. http://www.r-project.org/

43. Must, A., & Anderson, S. E. (2006). Body mass index in children and adolescents: considerations for population-based applications. International Journal of Obesity, 30(4), 590–594.

44. Maher, C. A., & Olds, T. S. (2011). Minutes, MET minutes, and METs: unpacking socio-economic gradients in physical activity in adolescents. Journal of Epidemiology & Community Health, 65(2), 160–165.

45. Brockman, R., Jago, R., & Fox, K. R. (2010). The contribution of active play to the physical activity of primary school children. Preventive Medicine, 51(2), 144–147.

46. Mooses, K., & Kull, M. (2019). The participation in organised sport doubles the odds of meeting physical activity recommendations in 7–12-year-old children. https://Doi.Org/10.1080/17461391.2019.1645887, 20(4), p563–569.

47. Bann, D., Scholes, S., Fluharty, M., & Shure, N. (2019). Adolescents’ physical activity: Cross-national comparisons of levels, distributions and disparities across 52 countries. International Journal of Behavioral Nutrition and Physical Activity, 16(1), 141.

48. Statistical Yearbook of Portugal 2002. (2003). Instituto Nacional De Estatistica.

49. Freitas, D., Maia, J., Beunen, G., Claessens, A., Thomis, M., Marques, A., Crespo, M., & Lefevre, J. (2007). Socio-economic status, growth, physical activity and fitness: The Madeira Growth Study. Annals of Human Biology, 34(1), 107–122.

50. Watson, A., Eliott, J., & Mehta, K. (2015). Perceived barriers and facilitators to participation in physical activity during the school lunch break for girls aged 12–13 years: http://Dx.Doi.Org/10.1177/1356336X14567545, 21(2), p257–271.

51. González, S. A., Aubert, S., Barnes, J. D., Larouche, R., & Tremblay, M. S. (2020). Profiles of Active Transportation among Children and Adolescents in the Global Matrix 3.0 Initiative: A 49-Country Comparison. International Journal of Environmental Research and Public Health, 17(16), 1–29.

52. Pucher, J., & Buehler, R. (2008). Making Cycling Irresistible: Lessons from The Netherlands, Denmark and Germany. Transport Reviews, 28(4), 495–528.

53. Claessens, S., & Atkinson, Q. (n.d.). The non-independence of nations and why it matters.

54. Mattocks, C., Ness, A., Leary, S., Tilling, K., Blair, S. N., Shield, J., Deere, K., Saunders, J., Kirkby, J., Smith, G. D., Wells, J., Wareham, N., Reilly, J., & Riddoch, C. (2008). Use of accelerometers in a large field-based study of children: protocols, design issues, and effects on precision. Journal of Physical Activity & Health, 5 Suppl 1(SUPPL. 1).

55. Crawford, D., Cleland, V., Timperio, A., Salmon, J., Andrianopoulos, N., Roberts, R., Giles-Corti, B., Baur, L., & Ball, K. (2010). The longitudinal influence of home and neighbourhood environments on children’s body mass index and physical activity over 5 years: the CLAN study. International Journal of Obesity 2010 34:7, 34(7), 1177–1187.

56. Riddoch, C., Edwards, D., Page, A., Froberg, K., Anderssen, S. A., Wedderkopp, N., Brage, S., Cooper, A. R., Sardinha, L. B., Harro, M., Klasson-Heggebø, L., van Mechelen, W., Boreham, C., Ekelund, U., & Andersen, L. B. (2005). The European Youth Heart Study— Cardiovascular Disease Risk Factors in Children: Rationale, Aims, Study Design, and Validation of Methods. Journal of Physical Activity and Health, 2(1), 115–129.

57. Jackson, M., Crawford, D., Campbell, K., & Salmon, J. (2008). Are parental concerns about children’s inactivity warranted, and are they associated with a supportive home environment? Research Quarterly for Exercise and Sport, 79(3), 274–282.

58. Salmon, J., Campbell, K. J., & Crawford, D. A. (2006). Television viewing habits associated with obesity risk factors: a survey of Melbourne schoolchildren. Medical Journal of Australia, 184(2), 64–67.

59. Zahner, L., Puder, J. J., Roth, R., Schmid, M., Guldimann, R., Pühse, U., Knöpfli, M., Braun-Fahrländer, C., Marti, B., & Kriemler, S. (2006). A school-based physical activity program to improve health and fitness in children aged 6-13 years (‘Kinder-Sportstudie KISS’): Study design of a randomized controlled trial [ISRCTN15360785]. BMC Public Health, 6(1), 1–12.

60. Komanduri, S., Jadhao, Y., Guduru, S. S., Cheriyath, P., & Wert, Y. (2017). Prevalence and risk factors of heart failure in the USA: NHANES 2013 – 2014 epidemiological follow-up study. Journal of Community Hospital Internal Medicine Perspectives, 7(1), 15–20.

61. Wolff-Hughes, D. L., Bassett, D. R., & Fitzhugh, E. C. (2014). Population-Referenced Percentiles for Waist-Worn Accelerometer-Derived Total Activity Counts in U.S. Youth: 2003 – 2006 NHANES. PLoS ONE, 9(12), e115915.

62. Victora, C. G., Hallal, P. C., Araújo, C. L. P., Menezes, A. M. B., Wells, J. C. K., & Barros, F. C. (2008). Cohort Profile: The 1993 Pelotas (Brazil) Birth Cohort Study. International Journal of Epidemiology, 37(4), 704–709.

