## Supplementary Data for "Gender differences in the distribution of children’s physical activity: evidence from nine countries"

### *Supplementary Information*

Table S 9: Association between gender and volumes of sedentary activity for the restricted model adjusted for gender, country, BMI (z-score) and parental education (as presented in the paper) and two further models, one additionally adjusted for season, one additionally adjusted for ethnicity... 47
Table S 10: Association between gender and volumes of light-intensity physical activity for the restricted model adjusted for gender, country, BMI (z-score) and parental education (as presented in the paper) and two further models, one additionally adjusted for season, one additionally adjusted for ethnicity........ 48
Table S 11: Association between gender and moderate activity as defined by Evenson cut points.. 50
Table S 12: Association between gender and vigorous activity as defined by Evenson cut points... 51

A complete markdown document for all analyses (Cleaning, analysis in the main article and those in the sensitivity analysis) are available here.

***Table S 1: Sample Characteristics, stratified by country. a = Data not collected in marked study. b = The response categories for mother and father education in EYHS Portugal did not include options for vocational training or distinguish those who started university / further education but did not complete it. It was, therefore, only possible to assign participants to two of the three categories for this harmonised variable (Up to and including completion of compulsory education (coded 0) / Completed undergraduate or postgraduate education (2))At the time of writing, one university offering degrees was in operation in Funchal, with a total student body of 2,898 individuals, which may represent restricted access for parents included in the Portuguese sample.[48] c = Detail on fathers' education was absent in the Brazilian (Pelotas) study***

| | **Overall** | **Australia** | **Brazil** | **Denmark** | **Estonia** | **Norway** | **Portugal** | **Switzerland** | **UK** | **USA** |
|---|---|---|---|---|---|---|---|---|---|---|
| **n** | 18980 | 2542 | 456 | 1636 | 659 | 387 | 1184 | 500 | 6696 | 4920 |
| **Gender (Boy) (%)** | 9176 (48.3) | 1210 (47.6) | 238 (52.2) | 734 (44.9) | 293 (44.5) | 197 (50.9) | 589 (49.7) | 242 (48.4) | 3204 (47.8) | 2469 (50.2) |
| **Age (mean (SD))** | 11.80 (2.71) | 9.62 (2.62) | 13.34 (0.31) | 12.34 (2.95) | 12.42 (3.00) | 9.68 (0.33) | 10.62 (2.61) | 9.31 (2.12) | 12.07 (0.78) | 12.84 (3.52) |
| **Height (mean (SD))** | 149.43 (15.35) | 137.40 (16.19) | 158.02 (8.40) | 153.08 (16.75) | 152.74 (17.50) | 139.20 (6.36) | 141.35 (13.66) | 136.16 (13.05) | 152.13 (8.42) | 153.36 (17.87) |
| **Weight (mean (SD))** | 45.33 (16.79) | 36.45 (13.07) | 51.10 (11.98) | 45.88 (15.78) | 44.66 (15.83) | 33.17 (5.87) | 38.66 (12.97) | 32.63 (9.81) | 44.93 (11.02) | 53.50 (22.37) |
| **Ethnicity (White) (%)** | 9408 (69.3) | 0 ( NaN)[a] | 303 (67.0) | 1096 (94.3) | 584 (97.7) | 306 (83.2) | 638 (98.0) | 0 ( NaN) | 5218 (96.2) | 1263 (25.7) |
| **Mothers Education (Beyond Compulsory) (%)** | 9004 (58.5) | 1134 (46.5) | 104 (22.8) | 1165 (79.2) | 418 (70.7) | 190 (53.1) | 31 ( 4.9)[b] | 314 (79.7) | 3691 (77.0) | 1957 (46.0) |
| **Fathers Education (Beyond Compulsory) (%)** | 7677 (61.0) | 1272 (60.3) | 0 ( NaN)[c] | 1110 (78.8) | 354 (65.4) | 189 (61.6) | 11 ( 1.8)[b] | 330 (86.6) | 3084 (72.9) | 1327 (44.2) |

### *Studies included in analysis*

***Table 6: Summary of studies included in ICAD employed in this study prior to data reduction.***

| Study | Country (Region) | Year of Recording | Number of Subjects Included | Device Used | Age Range (Years) |
|---|---|---|---|---|---|
| **ALSPAC** | United Kingdom (Bristol) | 2003 - 2007 | 6935 | Actigraph GT1M | 10 - 15 |
| **CLAN** | Australia (Melbourne) | 2001, 2004, 2006 | 1202 | Actigraph GT1M | 5 - 18 |
| **EYHS** | Denmark (Odense) | 1997 – 1998, 2003 - 2004 | 1814 | Actigraph GT1M & GT3X | 8 – 18 |
| **EYHS** | Estonia (Tartu) | 1998 - 1999 | 662 | Actigraph AM 7164 | 8 – 17 |
| **EYHS** | Norway (Oslo) | 1999 - 2000 | 398 | Actigraph AM 7164 | 9 - 10 |
| **EYHS** | Portugal (Madeira) | 1999 - 2000 | 1256 | Actigraph GT1M | 8 – 18 |
| **HEAPS** | Australia (Melbourne) | 2002 – 2003, 2006 | 1453 | Actigraph GT1M | 4 – 16 |
| **KISS** | Switzerland (Aargau and Basel) | 2005 - 2006 | 532 | Actigraph GT1M | 6 - 14 |
| **NHANES** | USA (National) | 2003 – 2004, 2005 - 2006 | 5284 | Actigraph AM-7164 | 6 – 18 |
| **Pelotas** | Brazil (Pelotas) | 2006 - 2007 | 457 | Actigraph GT1M | 13 - 14 |

#### *1.1.1.1 Avon Longitudinal Study of Parents and Children (ALSPAC) – UK*

The Avon Longitudinal Study of Parents and Children is a longitudinal study conducted in the Bristol area. The study initially recruited 14,541 pregnant women who had expected deliveries between April 1991 and December 2002.[36,37] This resulted in 14,062 live births, with a further 913 participants joining later (these were from mothers who were eligible at the initial recruitment phase but did not join the study until a later point). In interviews between 2003 and 2007 (approximate ages of 14 to 16years) individuals were invited to wear a waist-worn accelerometer for 7 consecutive days.[54] Of the initial study sample, 6060 individuals contributed valid activity data that was included in ICAD.[34]

##### *1.1.1.2 Children Living in Active Neighbourhoods Project (CLAN) – Australia*

The Children Living in Active Neighbourhoods Project is longitudinal study of 2,096 children from 19 schools in Melbourne, with three waves of data collection.[55] Individuals were initially recruited at ages 10-12 years, and followed up with two additional waves, three and five years later. At each wave physical activity data was recorded using a waist worn accelerometer. Of these 1,126 children provided valid physical activity data that could be incorporated into ICAD.[34]

##### *1.1.1.3 European Youth Heart Study (EYHS) - Denmark, Estonia, Norway, Portugal*

The European Youth Heart Study was conducted in Odense (Denmark), Tartu (Estonia), Oslo (Norway) and Madeira (Portugal), with one sweep in Estonia and Norway, and two sweeps in Denmark and Portugal.[56] Aiming to explore risk factors for cardio-vascular disease, accelerometery data was collected from participating individuals by waist worn accelerometers. At initial recruitment individuals were approximately aged 9 or 15years. From this a total of 1,308 individuals from Denmark, 662 individuals from Estonia, 391 individuals from Norway, and 1242 individuals from Portugal had data that could be incorporated into ICAD.[34]

##### *1.1.1.4 Healthy Eating and Play Study (HEAPS) – Australia*

The Healthy Eating and Play Study is a longitudinal study of children from Melbourne, aged either 5-to-6 years or 10-to-12 years of age at induction to the study and followed up 4 years later.[57,58] Individuals were requested to provide waist worn accelerometery, with 1,362 having complete data that could be contributed to ICAD.[34]

##### *1.1.1.5 Kinder-Sportstudie (KISS) – Switzerland*

The Kinder-Sportstudie is an intervention study designed to explore whether a school-based physical intervention could increase activity levels in a group of 6-to-13 year olds, conducted in the Aargau and Basel cantons of Switzerland.[59] Within these regions non-random sampling was employed, so as to be representative of the wider demographics of Switzerland. Participating individuals provided accelerometery data before and after the intervention by wearing waist-worn devices. A total of 433 individuals had complete data at baseline which could be contributed to ICAD.[34] While both pre and post intervention data is available, only the pre-test data is included in the present analyses.

##### *1.1.1.6 National Health and Nutrition Examination Survey (NHANES) – United States of America*

The National Health and Nutrition Examination Survey is a repeated cross-sectional study undertaken across the USA and aims to be broadly representative of the national population, with some oversampling of minority groups to ensure statistical power.[60] In the 2003 and the 2005 surveys, waist worn accelerometer data was collected from all consenting individuals aged 6 years or older.[61] Due to privacy concerns, the raw accelerometery files were not made available for inclusion in ICAD.[34] This means that the exact start date of the recording is not available. In the harmonisation process, the start date for each file was set to be between the 1st and 7th of January on the year of recording, with the date picked to match the day of the week on which recording started. Across the two survey years, a total of 5,174 individuals aged between 6 and 18 years had valid accelerometery data that could be included in ICAD.[34]

##### *1.1.1.7 Pelotas Study – Brazil*

The Pelotas Study is a multi-cohort longitudinal study that samples birth cohorts from the city of Pelotas in Brazil, repeated roughly every 10 years. The 1993 cohort consists of all the children born in the City of Pelotas during the calendar year (for whom their parents agreed to participation).[62] Objective physical activity data was collected from a subset of the sample in 2006 using waist worn devices. The subset was consistent of individuals who had been interviewed at every sweep to that point. From the subset collected in 2006, a total of 457 individuals had data that could be contributed to ICAD.[34]

### *Missingness by study and variable*

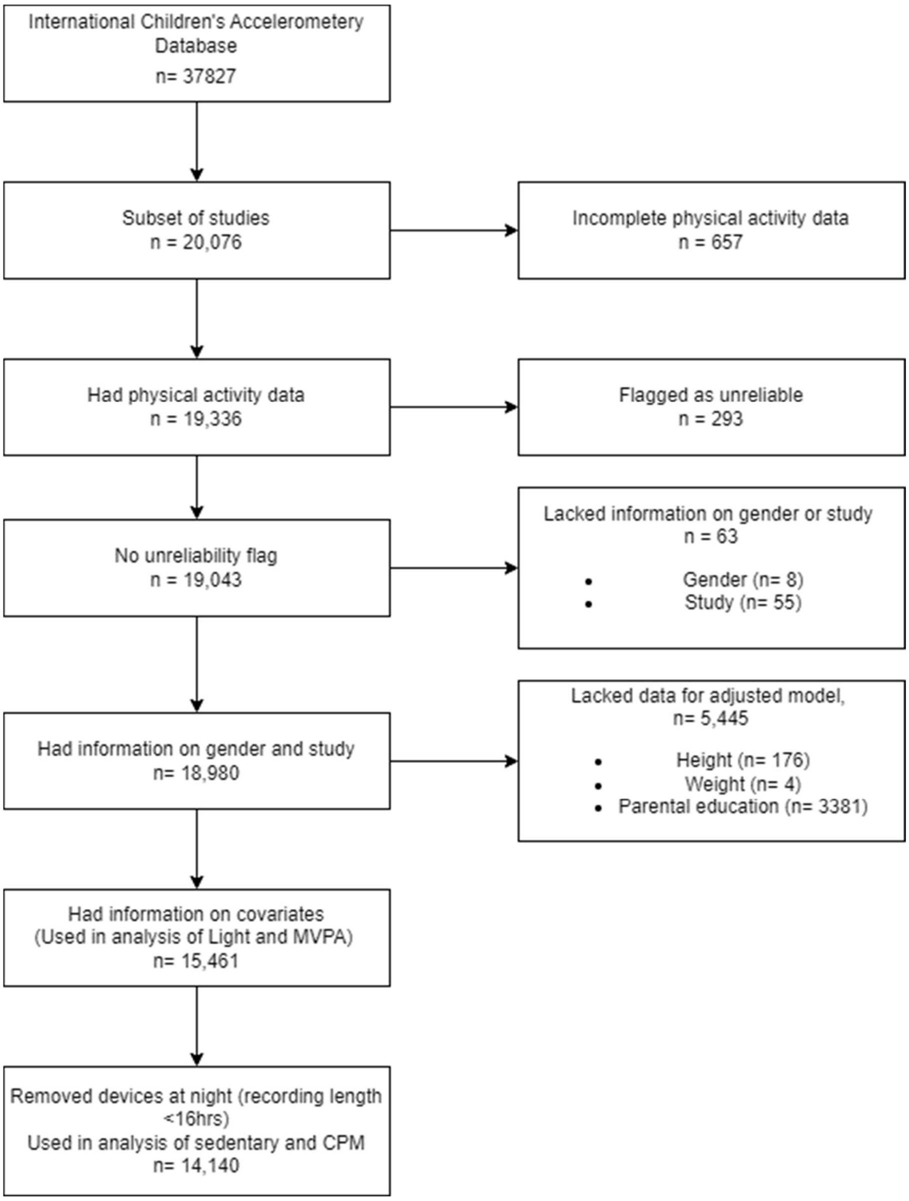

***Figure S 1 – Flow of missingness by core variables. Other restrictions were placed on day length and are detailed in the body text.***

***Table S 2 – Number of individuals with missing data by demographic variable and Gender***

| | **Overall** | **Girls** | **Boys** |
|---|---|---|---|
| **n** | 885 | 380 | 411 |
| **Age (mean (SD))** | 12.77 (3.30) | 12.70 (3.33) | 12.83 (3.29) |
| **Height (mean (SD))** | 154.03 (18.25) | 150.96 (16.36) | 156.78 (19.48) |

| | | | |
|---|---|---|---|
| **Weight (mean (SD))** | 50.83 (19.93) | 49.38 (19.58) | 52.17 (20.29) |
| **Ethnicity (white) (%)** | 287 (57.7) | 130 (56.0) | 157 (59.2) |
| **Country (%)** | | | |
| **Australia** | 93 (11.6) | 53 (13.9) | 40 ( 9.7) |
| **Brazil** | 1 ( 0.1) | 1 ( 0.3) | 0 ( 0.0) |
| **Denmark** | 85 (10.6) | 43 (11.3) | 42 (10.2) |
| **Estonia** | 2 ( 0.2) | 1 ( 0.3) | 1 ( 0.2) |
| **Norway** | 11 ( 1.4) | 2 ( 0.5) | 3 ( 0.7) |
| **Portugal** | 32 ( 4.0) | 15 ( 3.9) | 17 ( 4.1) |
| **Switzerland** | 32 ( 4.0) | 17 ( 4.5) | 15 ( 3.6) |
| **UK** | 236 (29.4) | 99 (26.1) | 132 (32.1) |
| **USA** | 310 (38.7) | 149 (39.2) | 161 (39.2) |
| **Mothers Education (Beyond Compulsory) (%)** | 280 (52.4) | 146 (54.1) | 131 (50.2) |
| **Fathers Education (Beyond Compulsory) (%)** | 231 (57.3) | 125 (62.8) | 103 (51.2) |
| **Reason for Exclusion (%)** | | | |
| **Missing Physical Activity Data** | 657 (74.2) | 300 (78.9) | 333 (81.0) |
| **Missing Country** | 55 ( 6.2) | 0 ( 0.0) | 0 ( 0.0) |
| **Missing Gender** | 8 ( 0.9) | 0 ( 0.0) | 0 ( 0.0) |

| | | | |
|---|---|---|---|
| **Flagged as unreliable** | 165 (18.6) | 80 (21.1) | 78 (19.0) |

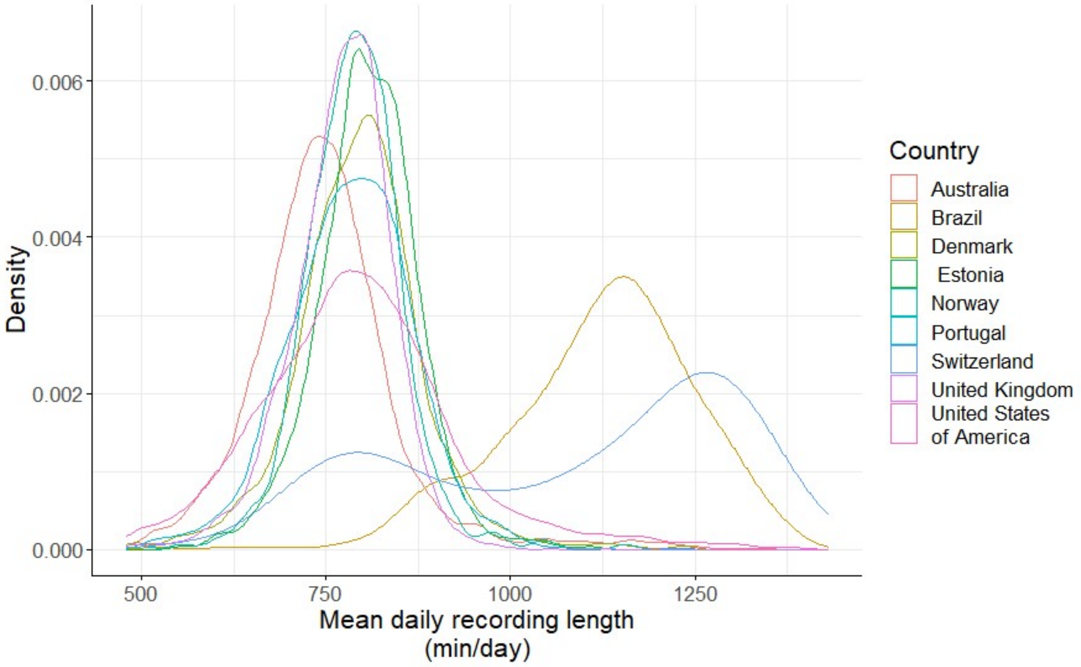

***Figure S 2: Density plot of mean recording length across nations of study.***

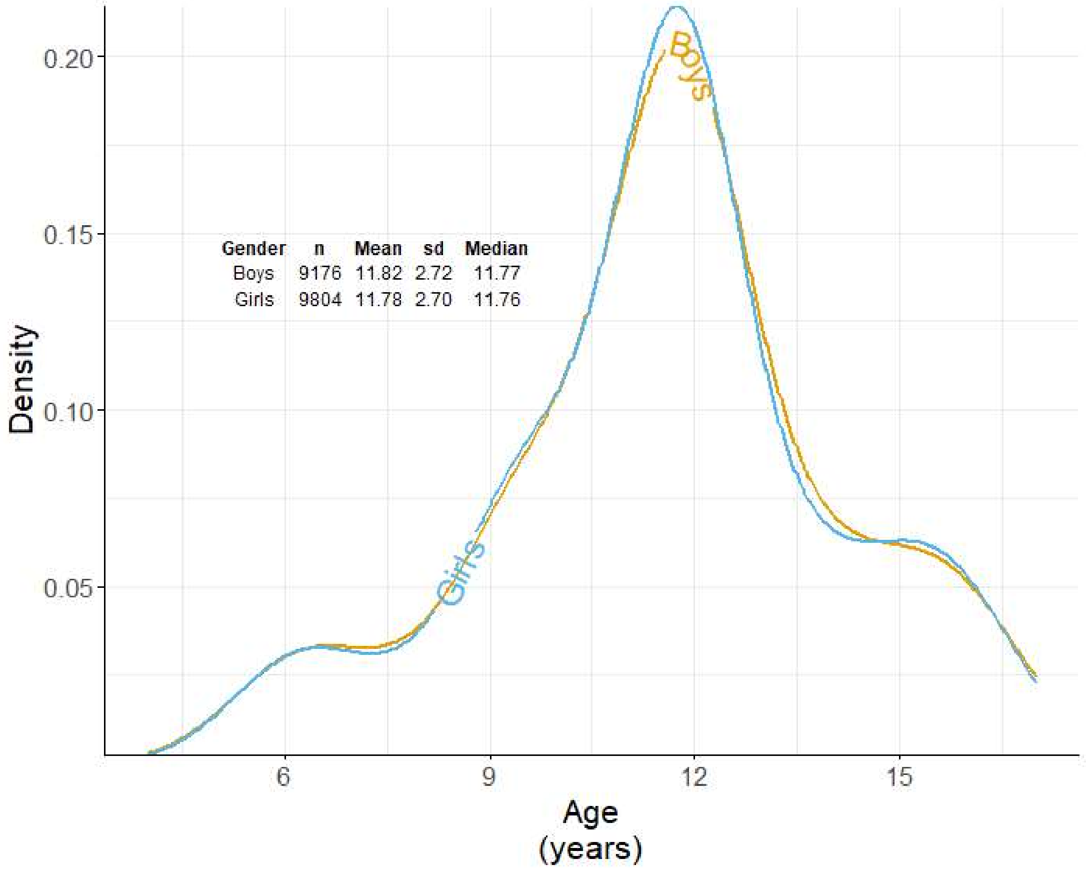

***Figure S 3: Density plot of age for girls and boys in the study.***

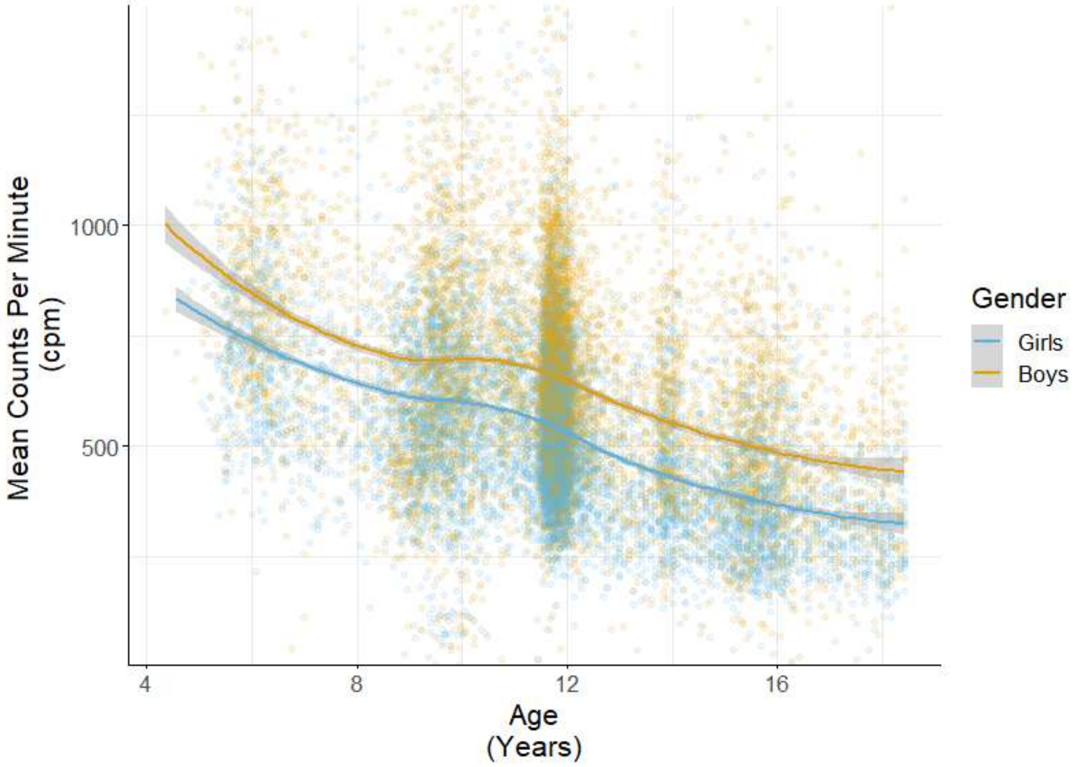

***Figure S 4: Mean counts per minute plotted against age including all individuals in the study. Line is best fit with 95% confidence interval marked in grey. Each colour represents one gender.***

***Gender differences in physical activity levels presented by country***

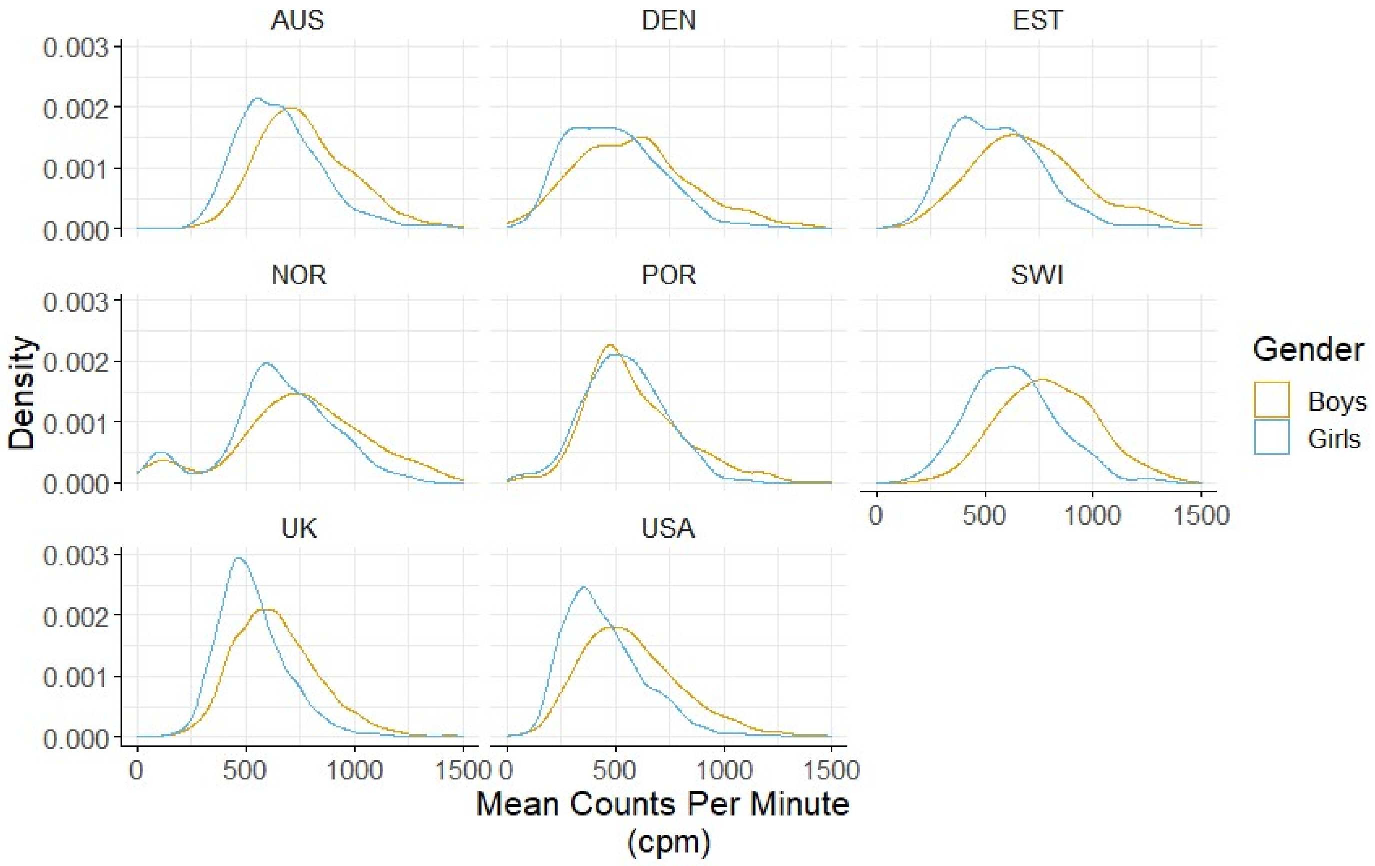

***Figure S 5: Density plot of mean counts per minute (cpm) by gender, presented by nation of study.***

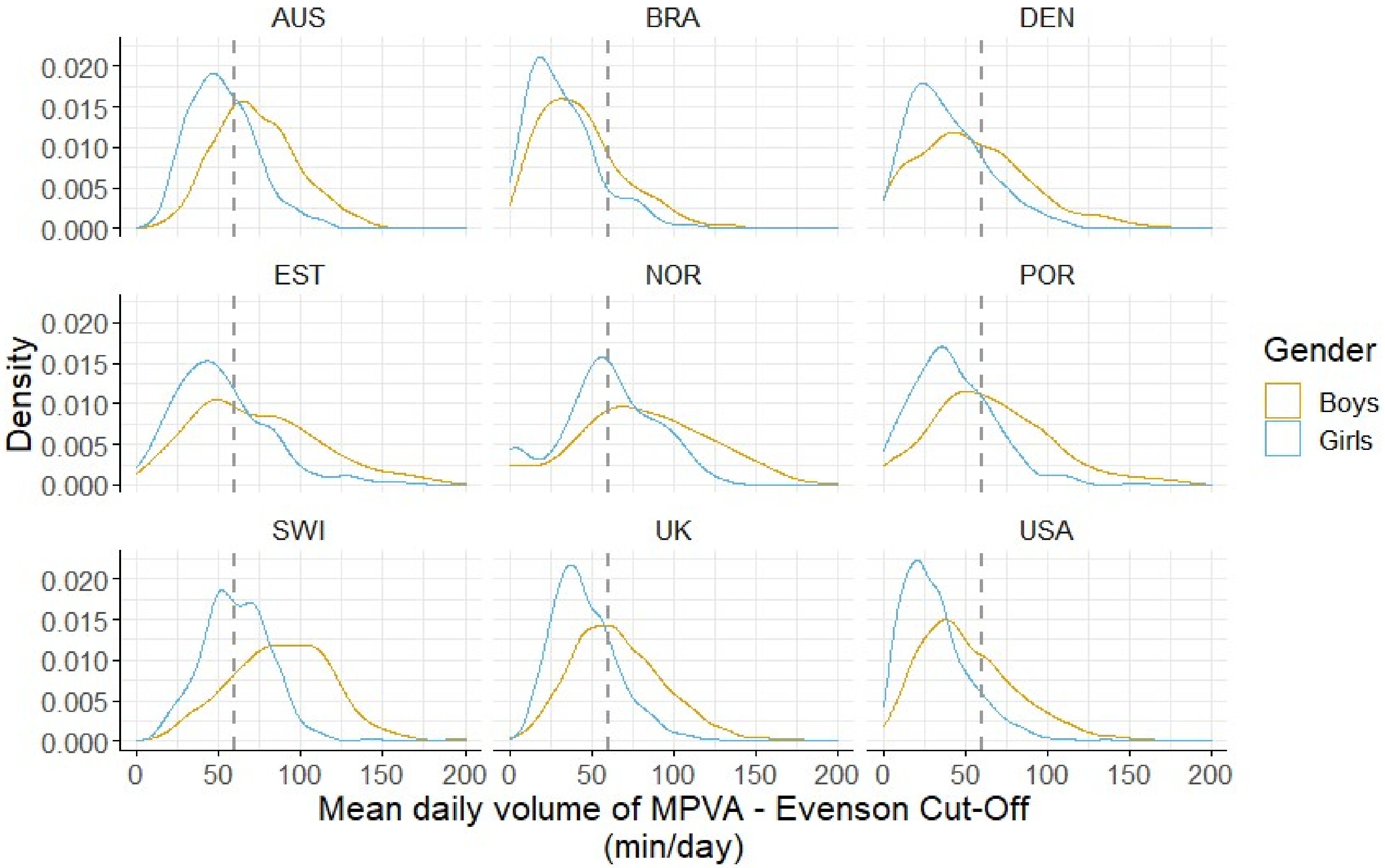

***Figure S 6: Density plot of mean daily minutes of moderate-to-vigorous physical activity (MVPA) by gender, presented by nation of study. Vertical dashed line represents 60 min/day of MVPA, WHO guidelines for recommend activity levels for a child aged 5-18.***

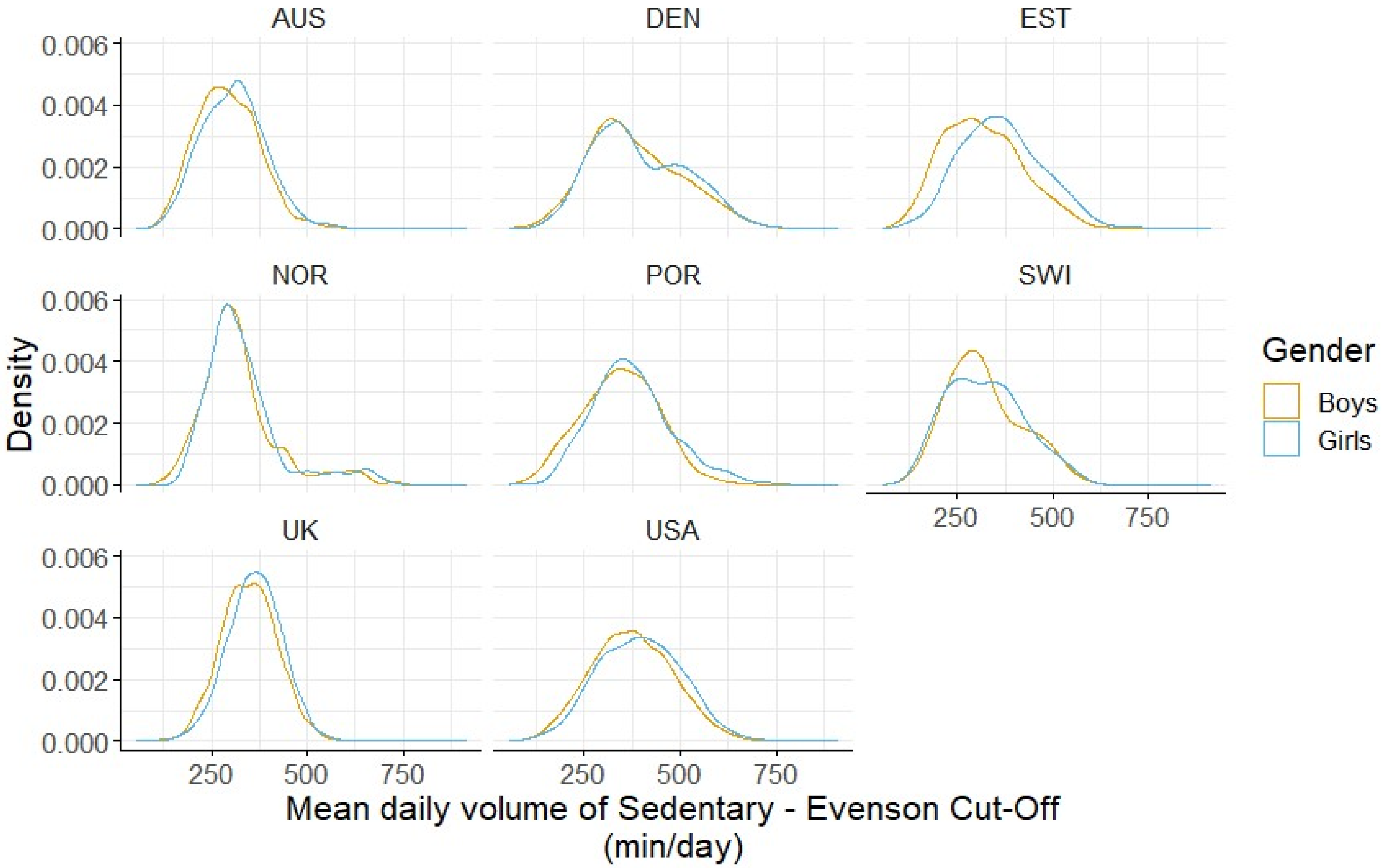

*Figure S 7: Density plot of mean daily minutes of sedentary activity by gender, presented by nation of study.*

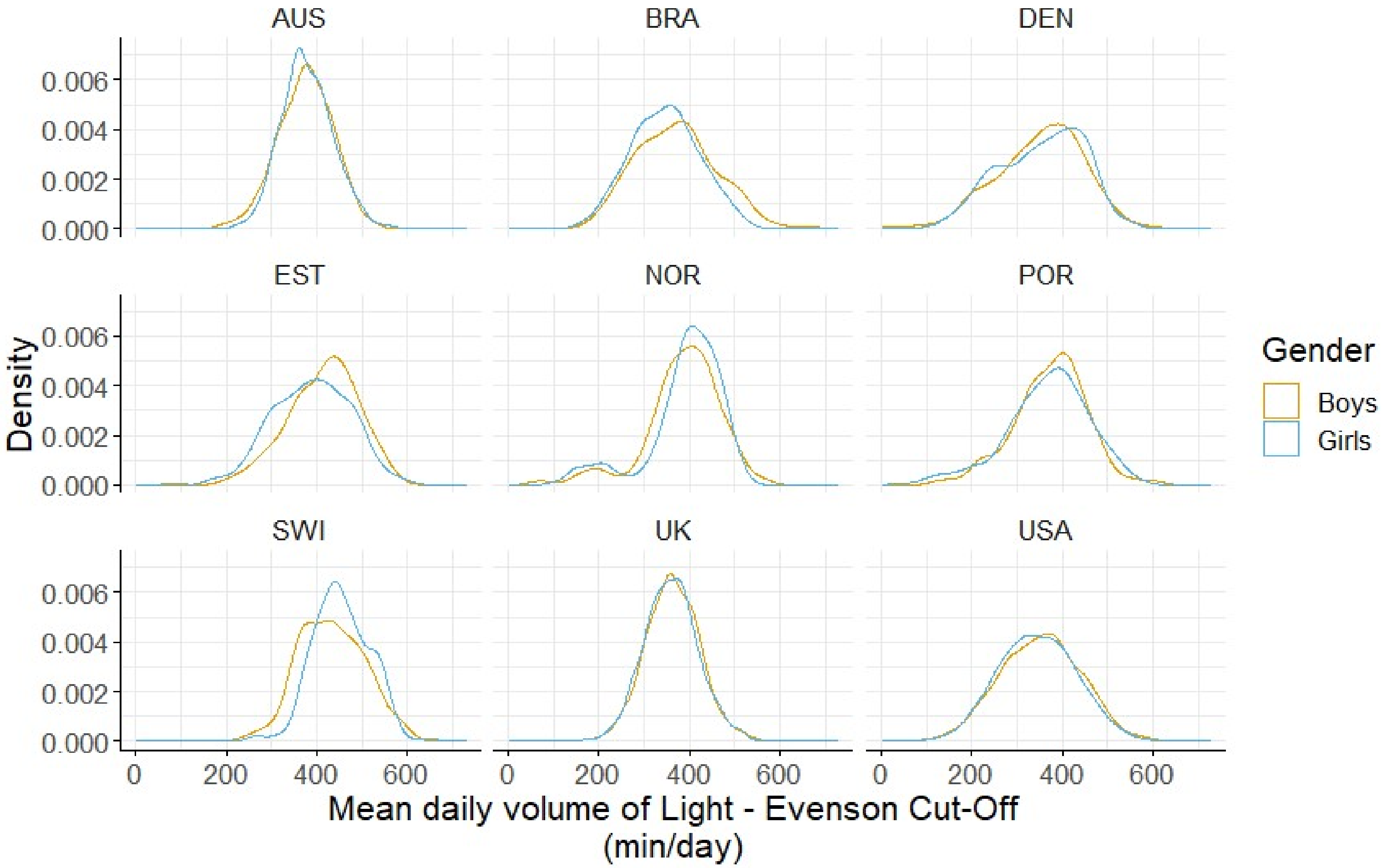

***Figure S 8: Density plot of mean daily minutes of light-intensity activity by gender, presented by nation of study.***

### *Sensitivity Analysis*

### *Unrestricted Samples*

***Table S 3: Association between gender and Counts Per Minute as defined by Evenson cut points for both restricted (as presented in the paper) and unrestricted sample sizes.*** *Differences in mean, variability and skew estimated by GAMLSS, a = samples after restriction for complete cases of for parental education, BMI, and country. NO: normal distribution. b = Skewness is estimated as the Box-Cox power (that is, the power required to transform the outcome to a normal distribution, values closer to 1 represent less skew). BCCG: Box-Cox Cole and Green distribution: SD: standard deviation. CoV: coefficient of variation. GAMLSS: Generalized Additive Models for Location, Scale and Shape. SE, standard error. * = p<0.05, ** = p<0.01, *** = p<0.001.*

| Gender | N (%) | NO distribution | | BCCG distribution | |
|---|---|---|---|---|---|
| | | **Mean** % Difference (SE) | **SD** % Difference (SE) | **Median** % Difference (SE) | **Skewness**[b] |
| **Girls** (ref) (Unrestricted Sample) | 9117 (52.3) | 530.4 | 198.0 | 511.4 | 0.48 |
| **Girls**[a] (Restricted Sample) | 7377 (52.2) | 532.91 | 200.32 | 513.31 | 0.48 |
| **Boys** (Unrestricted Sample) | 8387 (47.9) | 19.0 (0.56) *** | 16.9 (1.07) *** | 20.1 (0.61) *** | 0.15 (0.02) *** |
| **Boys**[a] (Restricted Sample) | 6763 (47.8) | 19.85 (0.62) *** | 16.42 (1.26) *** | 21.29 (0.67) *** | 0.19 (0.03) *** |

***Table S 4: Association between gender and volumes of moderate-to-vigorous physical activity as defined by Evenson cut points for both restricted (as presented in the paper) and unrestricted sample sizes.*** *Differences in mean, variability and skew estimated by GAMLSS, a = samples after restriction for complete cases of for parental education, BMI, and country. NO: normal distribution. b = Skewness is estimated as the Box-Cox power (that is, the power required to transform the outcome to a normal distribution, values closer to 1 represent less skew). BCCG: Box-Cox Cole and Green distribution: SD: standard deviation. CoV: coefficient of variation. GAMLSS: Generalized Additive Models for Location, Scale and Shape. SE, standard error. * = p<0.05, ** = p<0.01, *** = p<0.001.*

| **Gender** | **N (%)** | **NO distribution** | | **BCCG distribution** | |
|---|---|---|---|---|---|
| | | **Mean** % Difference (SE) | **SD** % Difference (SE) | **Median** % Difference (SE) | **Skewness**[b] |
| **Girls** (ref) (Unrestricted Sample) | 9802 (51.7) | 43.51 | 24.17 | 40.27 | 0.55 |
| **Girls**[a] (Restricted Sample) | 7998 (51.7) | 42.94 | 23.80 | 39.64 | 0.56 |
| **Boys** (Unrestricted Sample) | 9164 (48.3) | 35.33 (0.78) *** | 28.57 (1.02) *** | 37.04 (0.88) *** | 0.06 (0.02) ** |
| **Boys**[a] (Restricted Sample) | 7463 (48.3) | 37.64 (0.86) *** | 29.50 (1.13) *** | 39.76 (0.98) *** | 0.08 (0.02) ** |

***Table S 5: Association between gender and volumes of sedentary activity as defined by Evenson cut points for both restricted (as presented in the paper) and unrestricted sample sizes.*** *Differences in mean, variability and skew estimated by GAMLSS, a = samples after restriction for complete cases of for parental education, BMI, and country. NO: normal distribution. b = Skewness is estimated as the Box-Cox power (that is, the power required to transform the outcome to a normal distribution, values closer to 1 represent less skew). BCCG: Box-Cox Cole and Green distribution: SD: standard deviation. CoV: coefficient of variation. GAMLSS: Generalized Additive Models for Location, Scale and Shape. SE, standard error. * = p<0.05, ** = p<0.01, *** = p<0.001.*

| **Gender** | **N (%)** | **NO distribution** | | **BCCG distribution** | |
|---|---|---|---|---|---|
| | | **Mean** % Difference (SE) | **SD** % Difference (SE) | **Median** % Difference (SE) | **Skewness**[b] |
| **Girls** (ref) (Unrestricted Sample) | 9117 (52.1) | 368.9 | 98.7 | 363.1 | 0.57 |
| **Girls**[a] (Restricted Sample) | 7377 (52.2) | 365.86 | 99.39 | 359.64 | 0.54 |
| **Boys** (Unrestricted Sample) | 8387 (47.9) | -5.11 (0.41) *** | -1.00 (1.07) | -5.29 (0.45) *** | -0.01 (0.04) |
| **Boys**[a] (Restricted Sample) | 6763 (47.8) | -5.01 (0.46) *** | -1.34 (1.19) | -5.15 (0.51) *** | 0.01 (0.05) |

***Table S 6: Association between gender and volumes of light-intensity activity as defined by Evenson cut points for both restricted (as presented in the paper) and unrestricted sample sizes.*** *Differences in mean, variability and skew estimated by GAMLSS, a = samples after restriction for complete cases of for parental education, BMI, and country. NO: normal distribution. b = Skewness is estimated as the Box-Cox power (that is, the power required to transform the outcome to a normal distribution, values closer to 1 represent less skew). BCCG: Box-Cox Cole and Green distribution: SD: standard deviation. CoV: coefficient of variation. GAMLSS: Generalized Additive Models for Location, Scale and Shape. SE, standard error. * = p<0.05, ** = p<0.01, *** = p<0.001.*

| Gender | N (%) | NO distribution | | BCCG distribution | |
|---|---|---|---|---|---|
| | | **Mean** % Difference (SE) | **SD** % Difference (SE) | **Median** % Difference (SE) | **Skewness[b]** |
| **Girls** (ref) (Unrestricted Sample) | 9804 (48.3) | 358.6 | 78.3 | 361.1 | 1.29 |
| **Girls[a]** (Restricted Sample) | 7998 (51.7) | 361.93 | 77.73 | 364.43 | 1.29 |
| **Boys** (Unrestricted Sample) | 9176 (48.3) | 1.06 (0.32) *** | 2.02 (1.03) * | 1.12 (0.33) *** | 0.02 (0.05) |
| **Boys[a]** (Restricted Sample) | 7463 (48.3) | 0.82 (0.34) * | 2.36 (1.14) * | 0.90 (0.36) * | 0.03 (0.05) |

### *Adjustment for Season and Ethnicity*

***Table S 7: Association between gender and counts per minute for the restricted model adjusted for gender, country, BMI (z-score) and parental education (as presented in the paper) and two further models, one additionally adjusted for season, one additionally adjusted for ethnicity.*** *Differences in mean, variability and skew estimated by GAMLSS NO: normal distribution. b = Skewness is estimated as the Box-Cox power (that is, the power required to transform the outcome to a normal distribution, values closer to 1 represent less skew). BCCG: Box-Cox Cole and Green distribution: SD: standard deviation. CoV: coefficient of variation. GAMLSS: Generalized Additive Models for Location, Scale and Shape. SE, standard error. * = p<0.05, ** = p<0.01, *** = p<0.001.*

| Gender | N (%) | NO distribution | | BCCG distribution | |
|---|---|---|---|---|---|
| | | **Mean** % Difference (SE) | **SD** % Difference (SE) | **Median** % Difference (SE) | **Skewness**[b] |
| **Girls** (Adjusted Model) | 7377 (52.2) | 546.70 | 187.86 | 520.18 | 0.48 |
| **Girls** (Adjusted Model + Season) | 5359 (52.6) | 658.50 | 183.50 | 634.58 | 0.62 |
| **Girls** (Adjusted Model + Ethnicity) | 5750 (52.0) | 556.77 | 184.10 | 544.22 | 0.50 |
| **Boys** (Adjusted Model) | 7463 (48.3) | 37.70 (0.79) *** | 30.25 (1.14) *** | 40.49 (0.92) *** | 0.08 (0.02) *** |
| **Boys** (Adjusted Model + Season) | 4832 (47.4) | 18.70 (0.64) *** | 17.10 (1.40) *** | 19.62 (0.69) *** | 0.16 (0.04) *** |
| **Boys** (Adjusted Model + Ethnicity) | 5300 (48.0) | 20.58 (0.68) *** | 20.20 (1.35) *** | 21.69 (0.73) *** | 0.17 (0.03) *** |

***Table S 8: Association between gender and volumes of moderate to vigorous physical activity for the restricted model adjusted for gender, country, BMI (z-score) and parental education (as presented in the paper) and two further models, one additionally adjusted for season, one additionally adjusted for ethnicity.*** *Differences in mean, variability and skew estimated by GAMLSS NO: normal distribution. b = Skewness is estimated as the Box-Cox power (that is, the power required to transform the outcome to a normal distribution, values closer to 1 represent less skew). BCCG: Box-Cox Cole and Green distribution: SD: standard deviation. CoV: coefficient of variation. GAMLSS: Generalized Additive Models for Location, Scale and Shape. SE, standard error. * = p<0.05, ** = p<0.01, *** = p<0.001.*

| **Gender** | **N (%)** | **NO distribution** | | **BCCG distribution** | |
|---|---|---|---|---|---|
| | | **Mean** % Difference (SE) | **SD** % Difference (SE) | **Median** % Difference (SE) | **Skewness**[b] |
| **Girls** (Adjusted Model) | 7996 (51.7) | 53.02 | 22.36 | 47.27 | 0.53 |
| **Girls**[a] (Adjusted Model + Season) | 5822 (52.4) | 52.69 | 22.40 | 48.63 | 0.60 |
| **Girls** (Adjusted Model + Ethnicity) | 6165 (51.5) | 31.81 | 22.43 | 30.04 | 0.52 |
| **Boys** (Adjusted Model) | 7457 (48.3) | 37.59 (0.79) *** | 30.29 (1.14) *** | 40.34 (0.92) *** | 0.08 (0.02) *** |
| **Boys** (Adjusted Model + Season) | 5280 (47.6) | 35.58 (0.87) *** | 29.97 (1.34) *** | 36.54 (0.98) *** | 0.10 (0.03) *** |
| **Boys** (Adjusted Model + Ethnicity) | 5816 (48.5) | 39.38 (0.96) *** | 31.36 (1.30) *** | 42.24 (1.11) *** | 0.08 (0.02) *** |

***Table S 9: Association between gender and volumes of sedentary activity for the restricted model adjusted for gender, country, BMI (z-score) and parental education (as presented in the paper) and two further models, one additionally adjusted for season, one additionally adjusted for ethnicity.*** *Differences in mean, variability and skew estimated by GAMLSS NO: normal distribution. b = Skewness is estimated as the Box-Cox power (that is, the power required to transform the outcome to a normal distribution, values closer to 1 represent less skew). BCCG: Box-Cox Cole and Green distribution: SD: standard deviation. CoV: coefficient of variation. GAMLSS: Generalized Additive Models for Location, Scale and Shape. SE, standard error. * = p<0.05, ** = p<0.01, *** = p<0.001.*

| **Gender** | **N (%)** | **NO distribution** | | **BCCG distribution** | |
|---|---|---|---|---|---|
| | | **Mean** % Difference (SE) | **SD** % Difference (SE) | **Median** % Difference (SE) | **Skewness[b]** |
| **Girls** (Adjusted Model) | 7377 (52.2) | 354.35 | 94.67 | 346.52 | 0.70 |
| **Girls[a]** (Adjusted Model + Season) | 5359 (52.6) | 302.20 | 87.47 | 299.38 | 0.59 |
| **Girls** (Adjusted Model + Ethnicity) | 5750 (52.0) | 356.37 | 93.32 | 355.06 | 0.71 |
| **Boys** (Adjusted Model) | 6763 (47.8) | -5.04 (0.44) *** | -1.58 (1.19) | -5.18 (0.46) *** | -0.01 (0.05) |
| **Boys** (Adjusted Model + Season) | 4832 (47.2) | -5.03 (0.50) *** | -1.14 (1.40) | -5.02 (0.54) *** | 0.05 (0.06) |
| **Boys** (Adjusted Model + Ethnicity) | 5300 (48.0) | -4.88 (0.49) *** | -0.01 (1.35) | -4.92 (0.52) *** | 0.02 (0.05) |

***Table S 10: Association between gender and volumes of light-intensity physical activity for the restricted model adjusted for gender, country, BMI (z-score) and parental education (as presented in the paper) and two further models, one additionally adjusted for season, one additionally adjusted for ethnicity.*** *Differences in mean, variability and skew estimated by GAMLSS NO: normal distribution. b = Skewness is estimated as the Box-Cox power (that is, the power required to transform the outcome to a normal distribution, values closer to 1 represent less skew). BCCG: Box-Cox Cole and Green distribution: SD: standard deviation. CoV: coefficient of variation. GAMLSS: Generalized Additive Models for Location, Scale and Shape. SE, standard error. * = p<0.05, ** = p<0.01, *** = p<0.001.*

| Gender | N (%) | NO distribution | | BCCG distribution | |
|---|---|---|---|---|---|
| | | **Mean** % Difference (SE) | **SD** % Difference (SE) | **Median** % Difference (SE) | **Skewness[b]** |
| **Girls** (Adjusted Model) | 7998 (51.7) | 379.15 | 75.18 | 373.09 | 1.39 |
| **Girls[a]** (Adjusted Model + Season) | 5823 (52.4) | 374.28 | 69.72 | 372.54 | 1.56 |
| **Girls** (Adjusted Model + Ethnicity) | 6167 (51.4) | 360.92 | 76.59 | 366.84 | 1.37 |
| **Boys** (Adjusted Model) | 7463 (48.3) | 0.88 (0.33) ** | 3.26 (1.14) ** | 0.87 (0.35) * | -0.02 (0.06) |
| **Boys** (Adjusted Model + Season) | 5286 (47.6) | 0.45 (0.36) | 2.86 (1.34) * | 0.46 (0.37) | -0.08 (0.07) |
| **Boys** (Adjusted Model + Ethnicity) | 5822 (48 | 1.37 (0.39) *** | 3.47 (1.30) ** | 1.26 (0.41) ** | -0.02 (0.06) |

### *Separating MVPA into Moderate and Vigorous Intensity Activity*

#### *Moderate Intensity Physical Activity*

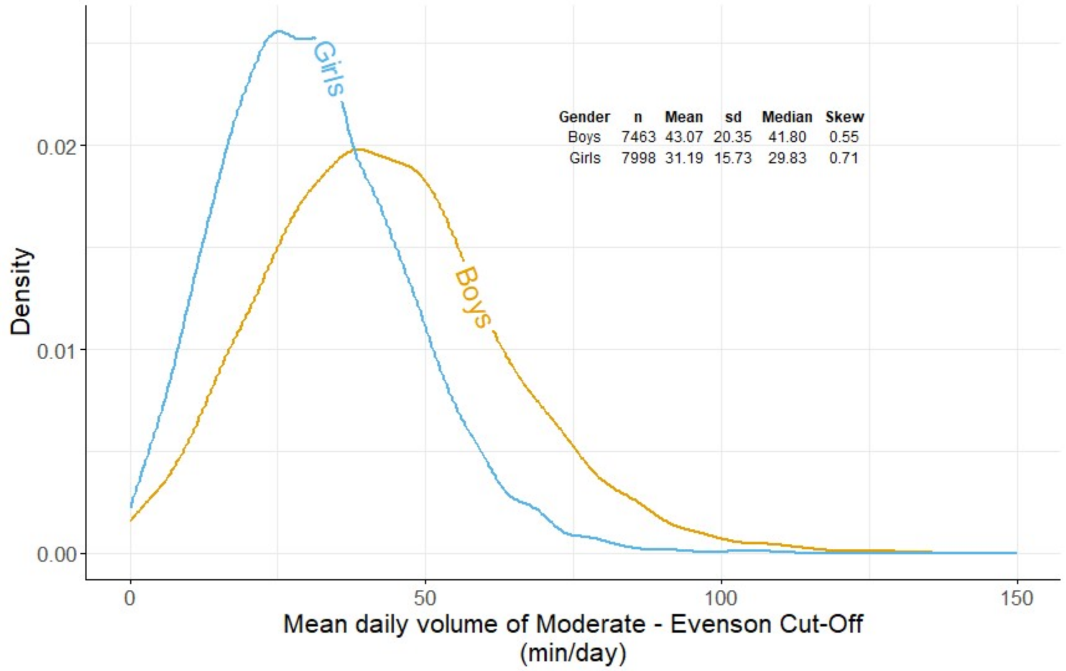

***Figure S 9: Density plot of moderate activity as defined by Evenson cut points. Each line representing a gender. Plot is muted at 150 min/day to show centre of distribution.***

***Table S 11: Association between gender and moderate activity as defined by Evenson cut points.*** *Differences in mean, variability and skew estimated by GAMLSS, n=, a = adjusted for parental education, BMI, and country. NO: normal distribution. b = Skewness is estimated as the Box-Cox power (that is, the power required to transform the outcome to a normal distribution). BCCG: Box-Cox Cole and Green distribution: SD: standard deviation. CoV: coefficient of variation. GAMLSS: Generalized Additive Models for Location, Scale and Shape. SE, standard error. * = p<0.05, ** = p<0.01, *** = p<0.001.*

| **Risk factor** | **N (%)** | **NO distribution** | | **BCCG distribution** | |
|---|---|---|---|---|---|
| | | **Mean** % Difference (SE) | **SD** % Difference (SE) | **Median** % Difference (SE) | **Skewness**[b] |
| **Girls** (ref) Unadjusted | 7998 (51.7) | 31.19 | 15.72 | 29.56 | 0.64 |
| **Boys** (Unadjusted Difference) | 7463 (48.3) | 32.27 (0.79) *** | 25.76 (1.14) *** | 33.59 (0.88) *** | 0.06 (0.02) ** |
| **Boys**[a] (Adjusted Difference) | 7463 (48.3) | 32.38 (0.76) *** | 26.54 (1.14) *** | 34.32 (0.84) *** | 0.07 (0.02) * |

*Vigorous Intensity Physical Activity*

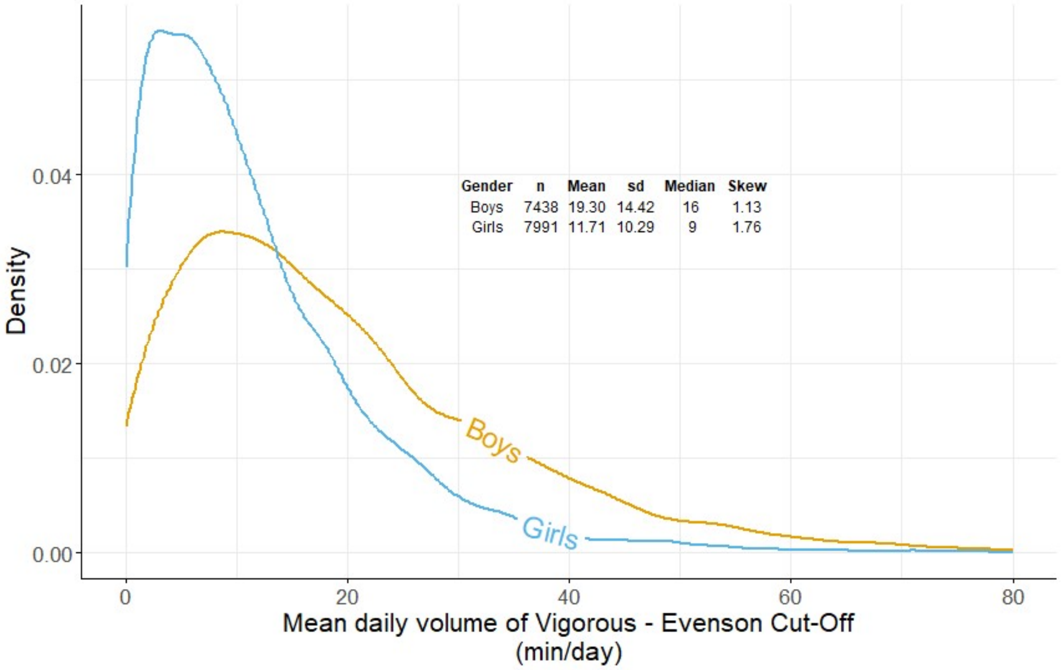

***Figure S 10: Density plot of vigorous activity as defined by Evenson cut points. Each line representing a gender. Plot is muted at 75 min/day to show centre of distribution.***

***Table S 12: Association between gender and vigorous activity as defined by Evenson cut points.*** *Differences in mean, variability and skew estimated by GAMLSS, n=, a = adjusted for parental education, BMI, and country. NO: normal distribution. SD: standard deviation. GAMLSS: Generalized Additive Models for Location, Scale and Shape. SE, standard error. * = p<0.05, ** = p<0.01, *** = p<0.001.*

| **Risk factor** | **N (%)** | **NO distribution** | |
|---|---|---|---|
| | | **Mean** % Difference (SE) | **SD** % Difference (SE) |
| **Girls** (ref) Unadjusted | 7991 (51.8) | 11.71 | 10.28 |
| **Boys** (Unadjusted Difference) | 7438 (48.2) | 49.93 (1.31) *** | 33.78 (1.14) *** |
| **Boys**[a] (Adjusted Difference) | 7438 (48.2) | 49.81 (1.22) *** | 33.66 (1.14) *** |
